# Small dried fish as an affordable source of key micronutrients in Madagascar: nutritional benefits and contamination risks

**DOI:** 10.64898/2026.08.28.26361595

**Authors:** Léono D. Todimazava, Maria J. Darias, Claire Mouquet-Rivier, Jamal Mahafina, Thomas Lamy

## Abstract

Micronutrient deficiencies are prevalent in Madagascar, where diets rely heavily on starchy staples and access to animal-source foods is limited. Small dried fish (SDF) are widely available, yet their nutritional value and health risks remain poorly documented. We combined market surveys, taxonomic identification, and micronutrient and heavy metal analyses of nine SDF types collected along National Road 7. The samples encompassed 33 fish families, were dominated by small pelagic species (Clupeidae and Engraulidae), and were appreciated by consumers. A daily portion (5 g for infants; 10 g for young children and women of childbearing age) contributed substantially to Recommended Nutrient Intakes (RNIs). Across samples and groups, SDF were rich (>30% of RNI) in selenium and, for infants and young children, in calcium. All samples were a source of (>15% of RNI), or rich in, phosphorus, whereas iron contributions were more variable but often substantial. Several samples exceeded 100% of RNIs for selenium, calcium, iron, or manganese in infants and young children, and some were also sources of magnesium and, less frequently, zinc. Vitamin A was absent from sun-dried samples but detected in a smoked freshwater type. Heavy metal concentrations varied markedly, and portions of several types led to estimated exposures to inorganic arsenic or cadmium exceeding reference values, whereas freshwater species and some pelagic types showed a more favorable nutrition-risk balance. Overall, SDF are affordable, nutrient-dense foods with strong potential to alleviate micronutrient deficiencies in Madagascar, while highlighting the need for type-specific guidance to balance nutritional benefits and contamination risks.

## 1. Introduction

Malnutrition is a global issue impacting over 2.3 billion people worldwide (FAO et al., 2023a). Micronutrient deficiencies, *i.e.*, deficiencies in vitamins and minerals commonly referred to as hidden hunger, are among the most prevalent forms of malnutrition (Tulchinsky, 2010), with young children and women of childbearing age being particularly vulnerable (Farias et al., 2020). For instance, anemia is a major health issue causing fatigue and weakness, especially in women and children (Cappellini et al., 2020), and can result from deficiencies in iron or several vitamins. More broadly, micronutrient deficiencies can lead to permanent damage, weaken the immune system, and increase morbidity and mortality (ACF, 2024). They are major challenges, especially in low- and middle-income countries (LMICs) (FAO et al., 2022), where the prevalence of malnutrition remains critical.

Micronutrient deficiencies are caused by limited dietary diversity, inadequate intake of nutrient-rich foods (Thompson and Amoroso, 2011; Passarelli et al., 2024), and low nutrient bioavailability (Kiani et al., 2022). Currently, over 3.1 billion people cannot afford a healthy diet (FAO, 2024). In LMICs, nutritious diets remain inaccessible to large segments of the population (FAO et al., 2023b). In sub-Saharan Africa, for instance, 72% of the population lacks access to healthy food due to poverty (FAO et al., 2024), and more than 821 million Africans experienced severe or moderate food insecurity in 2022. As a result, food access and affordability remain critical barriers to achieving Sustainable Development Goal 2: Zero Hunger.

Recent research highlights the importance of aquatic foods, including animals, plants, and algae harvested and cultured in freshwater, brackish, or marine systems, as essential sources of fatty acids and key micronutrients such as iron, calcium, zinc, and vitamins A, D, and B12 (Golden et al., 2016, 2021; Hicks et al., 2019). In LMICs, small-scale fisheries are an essential pillar of coastal communities’ economies and livelihoods (Basurto et al., 2025). Since these fisheries harvest a diverse range of aquatic species, they provide key micronutrients that can help alleviate food and nutrition insecurity. However, aquatic foods are not equally accessible or affordable (FAO et al., 2023a). While they are widely available in coastal areas where small-scale fisheries operate, challenges related to transportation and preservation often limit access to fresh aquatic foods in poor inland rural areas. Ensuring access to nutritious and affordable aquatic foods in such regions is crucial, as they provide vitamins and minerals often lacking in other food sources.

Processed fish, commonly preserved through sun-drying or smoking, are of particular importance (Belton et al., 2022), as they offer two key advantages: they are typically consumed whole, including nutrient-rich parts such as heads and viscera (Byrd et al., 2021), and processing extends shelf life and facilitates long-distance transport, potentially supplying critical micronutrients to inland rural areas. In sub-Saharan Africa, for instance, small dried fish (SDF) are frequently traded up to 1,000 km from their fishing grounds (Kawarazuka and Béné, 2011). SDF are generally defined as small fish (<12 cm) preserved using traditional methods such as sun-drying or smoking, which reduce moisture content and enable prolonged storage at room temperature without packaging while limiting spoilage. Among SDF, small pelagic species are of particular interest, as they constitute a major catch in LMICs and provide high levels of micronutrients (Robinson et al., 2022). Yet, SDF also encompass a wide range of freshwater and coastal species. Research has often focused on preservation methods and value chains, leaving the contribution of SDF to food security insufficiently documented (Beveridge et al., 2013; Bogard et al., 2015; Belton et al., 2022).

Madagascar, one of the world’s poorest countries, has 77% of its population living below the extreme poverty line and is among the six countries where hunger persists at an alarming level (World Bank, 2022). In 2024, Madagascar ranked 124^th^ of 127 countries in the Global Hunger Index (Rao et al., 2024). More than 18 million people experienced moderate or severe food insecurity in 2021 (FAO et al., 2023b). Diets are typically low in protein and micronutrients, relying mainly on starchy staples such as rice, cassava, and sweet potato. Iron and vitamin A deficiencies are widespread, with nearly half of children aged 6–59 months and over one-quarter of women aged 15–49 suffering from anemia (INSTAT and ICF, 2022) (**Supplementary Table 1**). Recent evidence also points to deficiencies in calcium, iron, selenium, iodine, and vitamins B1, B3, B12, D3, and E (Unicef, 2023; Passarelli et al., 2024; Ministère de la Santé Publique de Madagascar et al., 2025).

Fisheries are a vital component of Madagascar’s economy, with production estimated at 154,978 metric tons (MAEP, 2023). They provide a source of food and income for over a million people (MRHP, 2012), yet the average annual fish consumption is only 4 kg per capita (FAO, 2024), far below the 15 kg per capita recommended by the European Food Safety Authority (EFSA, 2014). Proximity to fishing grounds and affordability likely explain the low levels of fish consumption. However, processed aquatic foods, particularly SDF, are far more affordable for many Malagasy households. SDF are widely available in both coastal and inland markets and include a broad range of small pelagic, freshwater, and coastal fish species. Small pelagic fish are largely supplied by well-established offshore fisheries, especially in western Madagascar, while resource scarcity and poverty drive mosquito net fishing (Short et al., 2018) in coastal lagoons, lakes, and rivers, increasing the supply of small coastal and freshwater fish, including juveniles (Raharinaivo et al., 2020). Despite their widespread availability, no study has yet assessed the affordability and nutritional composition of SDF in Madagascar. This represents an important gap, given the ecological implications of harvesting small fish and their importance for food security.

This study aims to address this gap. Specifically, it investigates four questions: are SDF widespread in Madagascar? Are they affordable and appreciated by the Malagasy population? To what extent can they contribute to Recommended Nutrient Intakes (RNIs) for infants, young children, and women of childbearing age? Does SDF consumption pose risks of heavy metal exposure? To answer these questions, we conducted market surveys and taxonomic identification of SDF. We then analyzed nine representative types, covering coastal, pelagic, and freshwater species, for vitamin A, key minerals, and heavy metals in order to evaluate both their nutritional value and potential health risks.

## 2. Materials and methods

### 2.1. Market survey and analysis of small dried fish types

From February to August 2022, we visited 16 markets across 12 cities along National Road 7 (NR7) (**Supplementary Table 2**), which connects the coastal city of Toliara in southwest Madagascar to Antananarivo in the central region. As the total number of SDF vendors varied across markets, ranging from just four in Ilakaka to 92 in Antananarivo, we randomly selected a subset of vendors in the seven largest markets, representing 10–70% of all vendors. In total, 112 SDF vendors were surveyed, with four to 12 vendors per market. The survey consisted of three steps. First, we recorded all the different types of SDF sold by each vendor. A type of SDF was defined as a set of specimens smaller than 12 cm in length and sharing a unique indigenous (Malagasy) name (**Fig. 1**). Each vendor typically sold several batches of SDF, and the Malagasy name of each batch was obtained from the vendors. Second, we carried out semi-structured interviews with vendors to ask whether each type was considered an appreciated food item. Finally, we asked vendors and customers about SDF consumption habits, including common cooking methods and typical side dishes eaten with each type. In total, 389 distinct batches of SDF were recorded across all 112 vendors (**Supplementary Tables 2 and 3**).

**Fig. 1.**
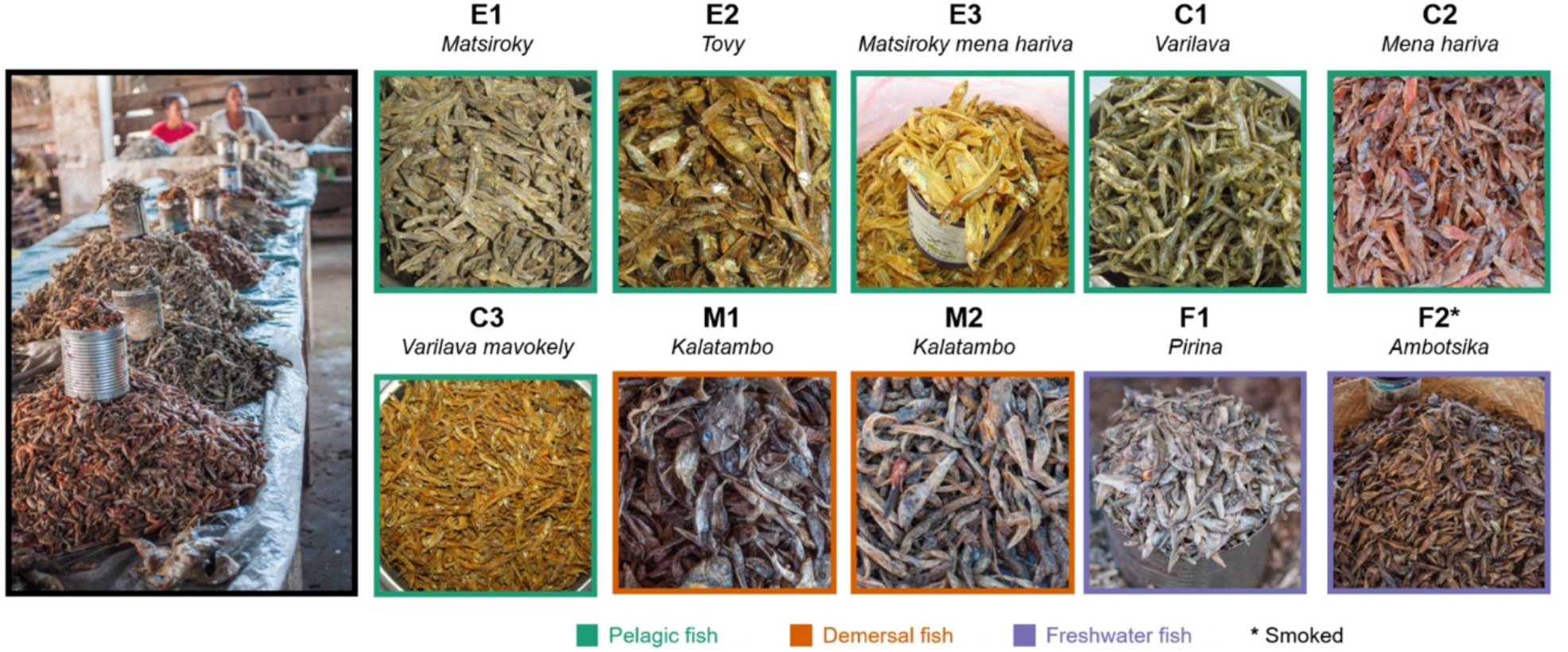
The ten samples of small dried fish (SDF) analyzed in this study. These samples represent nine distinct types, with their Malagasy names listed below each sample code: E = Engraulidae, C = Clupeidae, M = mixed fish, and F = freshwater fish. The photo on the left illustrates how SDF are sold in batches at the *Bazary Be* market in Sakaraha. The small cans placed on top of each batch, known locally as *Kapoaka*, serve as the standard sales unit for SDF

Following this large-scale survey, we focused on the nine most common SDF types (**Fig. 1**), which accounted for 275 of the 389 surveyed batches. They were grouped into three main categories: small pelagic fish, mixed small coastal fish, and freshwater fish, each referred to by its Malagasy name. The small pelagic fish category included six types: *Matsiroky*, *Tovy, Matsiroky mena hariva, Varilava*, *Mena hariva*, and *Varilava mavokely* (**Fig. 1**). The mixed small coastal fish category, known as *Kalatambo*, was sampled twice, resulting in a total of ten analyzed samples. *Kalatambo* comprised a variety of small demersal fish species, likely dominated by juvenile individuals. Lastly, the freshwater fish category included two types: *Pirina* and *Ambotsika*. Eight of these types were sun-dried, while *Ambotsika* was smoked (**Table 1**).

**Table 1.**
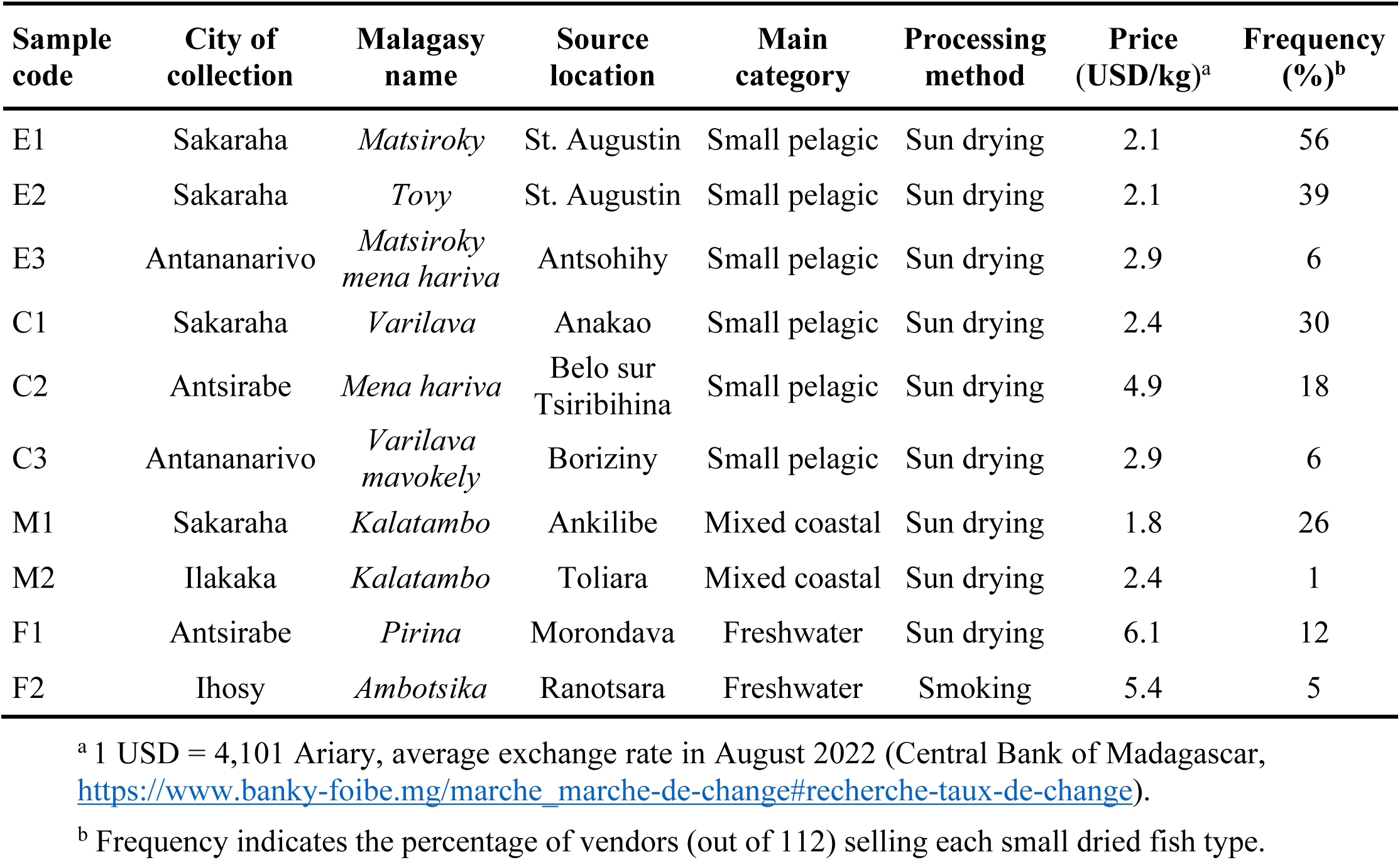
Main characteristics of the ten small dried fish samples analyzed.

### 2.2. Analysis of preferences and consumption patterns of small dried fish

We computed an appreciation rate based on survey responses for the 275 batches representing the nine focal SDF types. This score was calculated as the percentage of times a batch was considered an appreciated food item by vendors or consumers, obtained as the ratio of positive responses to total responses, multiplied by 100. In addition, we identified the most common cooking methods by calculating their relative frequencies for each SDF type. The same approach was applied to determine the relative frequencies of the most common side dishes.

### 2.3. Sampling of small dried fish

Ten samples, representing the nine focal SDF types, were collected in August 2022 from five inland cities located along the RN7 road from south to north: Sakaraha, Ilakaka, Ihosy, Antsirabe, and Antananarivo (**Table 1**). For each of the ten samples, one portion of the sales unit was first collected for taxonomic identification. This portion, averaging 60 g (range: 51– 74 g; **Table 2**), corresponded to the size of a metallic can locally known as a *Kapoaka,* which is traditionally used by vendors (**Fig. 1**). For nutritional analysis, we collected and stored 9–14 *Kapoaka* (∼700 g) of each SDF type in plastic bags. The price of one *Kapoaka* was recorded and converted to cost per kilogram.

**Table 2.**
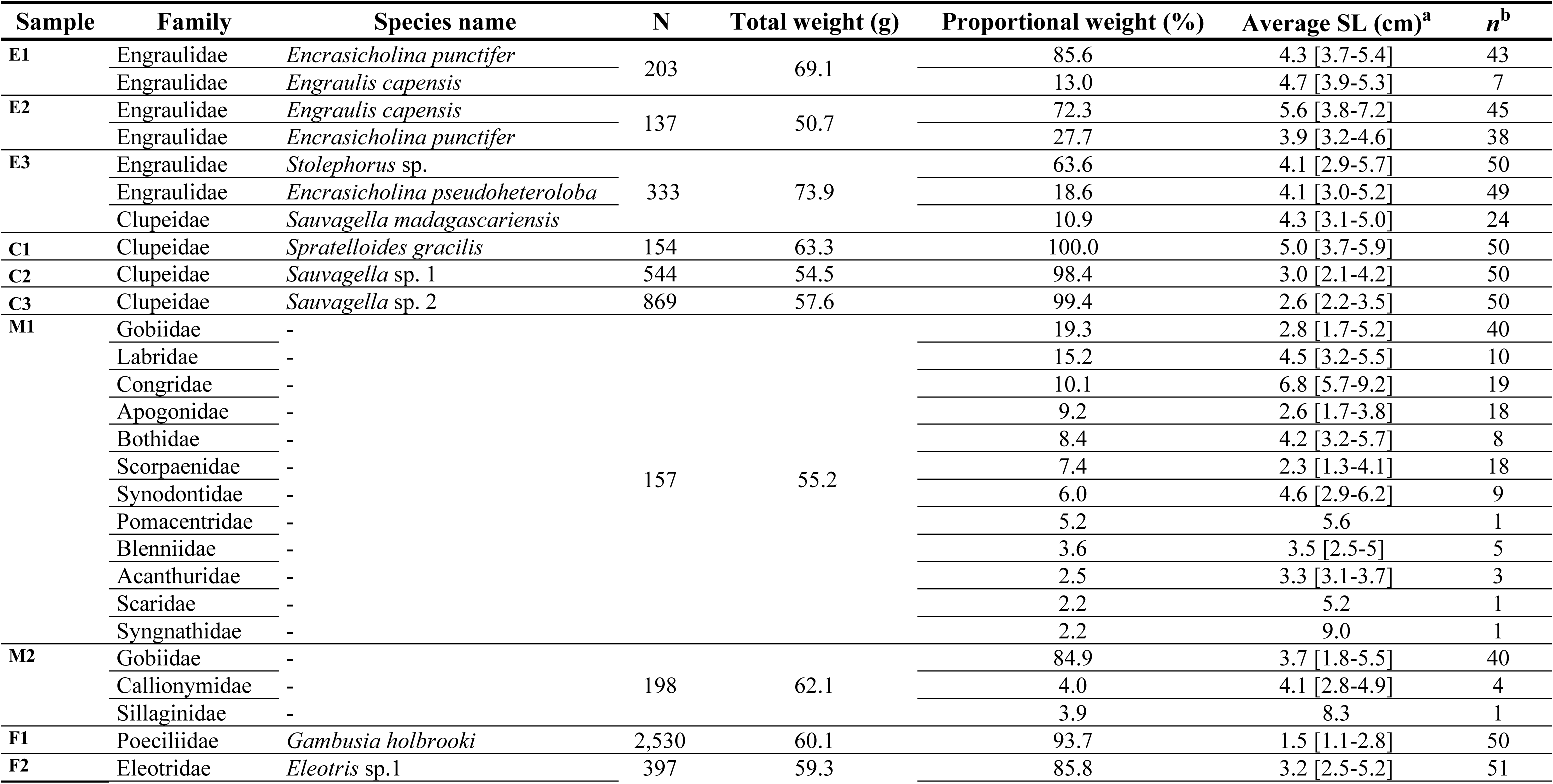
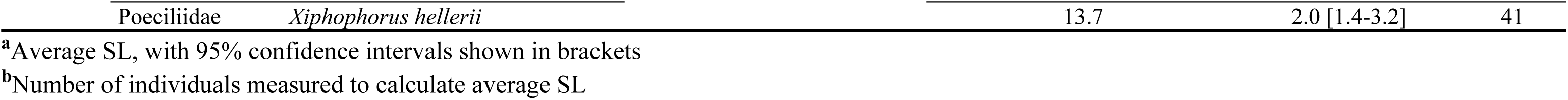
Taxonomic identification and size of the sampled small dried fish. For each sample, the family and scientific name of the main morpho-species are indicated, along with the number of individuals (N), total sample weight, proportional weight, and average standard length (SL). Morpho-species contributing less than 2% of the total biomass were excluded. Coastal fish taxa (M1 and M2) were classified at the family level due to difficulties in taxonomic identification.

### 2.4. Taxonomic identification and length measurement

For each sample, the taxonomic identity of all individuals was determined following the approach described in Raharinaivo et al. (2020). Briefly, all individuals were counted and sorted by morpho-species, defined as a set of specimens sharing similar morphological characteristics. Each fish was weighed, and up to 50 individuals per morpho-species were photographed. Standard length (SL) was measured from the images using ImageJ software (Schneider et al., 2012). Each morpho-species was identified to the lowest possible taxonomic level and cross-checked against a local checklist (Fricke et al., 2018).

Drying alters key morphological characters (e.g., fin rays, coloration), which may reduce identification accuracy and lead to misidentification. To resolve the taxonomy of certain morpho-species, we extracted DNA from 36 caudal fin fragments representing 16 morpho-species. Each fin fragment was stored in a 1.5 ml microtube with silica gel and frozen at –20 °C prior to DNA extraction. Total genomic DNA was extracted using the PureLink™ Genomic DNA Mini Kit (Thermo Fisher Scientific, Waltham, MA, USA), following the manufacturer’s instructions. PCR amplification and sequencing of the mitochondrial 12S gene were performed using primer pair XRMUPheF1 (5′YAAAGCATAMCRCTGAAGATG3′; Keleman et al., 2025) and teleo_R (5′CTTCCGGTACACTTACCATG3′; Valentini et al., 2016). Amplification was carried out in a total volume of 40 μl, containing 20 μl of DreamTaq™ Hot Start Master Mix (Thermo Fisher Scientific, Waltham, MA, USA), 0.6 μl of 3 μM forward and reverse primers mix, 0.8 μl of BSA 2%, and 2 μl of DNA template. The PCR conditions consisted of an initial denaturation at 92 °C for 3 min, followed by 35 cycles of denaturation at 92 °C for 45 s, annealing at 52 °C for 45 s, and elongation at 72 °C for 1 min, with a final extension at 72 °C for 5 min. Unidirectional Sanger sequencing was performed by Genoscreen (www.genoscreen.fr) and the nucleotide sequences were manually verified and edited using 4Peaks. The taxonomic identify of the amplified DNA sequences was evaluated using BLAST searches in the National Center for Biotechnology Information (NCBI) GenBank database (https://blast.ncbi.nlm.nih.gov/Blast.cgi). In addition, each DNA sequence was queried against a locally assembled 12S database (Volanandiana et al., 2025). Taxonomic assignment was based on BLAST best-hit similarities: to the species level for hits of 99–100%, to the genus level for hits of 96–99%, and to the family level for hits of 91–96%.

### 2.5. Biochemical analyses

Six 100 g replicates from each sample were finely ground using a Saisho blender. The powdered samples were stored in airtight bags with silica gel and kept at room temperature. Three replicates were used to quantify vitamin A, while the remaining three were used to quantify 10 minerals (calcium [Ca], iron [Fe], zinc [Zn], potassium [K], magnesium [Mg], sodium [Na], phosphorus [P], manganese [Mn], copper [Cu], selenium [Se]) and four heavy metals (cadmium [Cd], lead [Pb], mercury [Hg], arsenic [As]).

#### 2.5.1. Vitamin A

Vitamin A was measured in the form of retinol (including retinyl esters: retinyl acetate and retinyl palmitate) by Microchem Lab Services (Pty) Ltd (Cape Town, South Africa). Samples were saponified in a basic solution of ethanol and water, neutralized, and diluted, converting fats into fatty acids and retinyl esters. Quantification was performed by high-performance liquid chromatography (HPLC) with UV detection at 313 nm or 328 nm. Vitamin concentrations were calculated by comparing the peak areas of the analytes in the samples with those of calibration standards.

#### 2.5.2. Minerals and heavy metals

Minerals and heavy metals were analyzed by the Central Analytical Facilities of Stellenbosch University (South Africa). Samples were digested using a CEM MARS-5 microwave digester (CEM Corporation, Matthews, NC, USA) with concentrated ultrapure HNO3 and HCl. Trace elements such as Fe, Zn, Cu, Mn, As, Pb, Cd, and Hg were then quantified with an Agilent 7900 quadrupole ICP-MS (Agilent Technologies, Santa Clara, CA, USA). All analytes were measured in He collision mode, while H2 reaction gas was applied for Se. The elements Na, Mg, K, Ca, and P were analyzed using a Thermo Scientific iCAP 6200 ICP-AES (Thermo Fisher Scientific, Waltham, MA, USA) equipped with a charge-injection device solid-state detector. Calibration was performed with standards prepared in 2% HNO3 from multi-element stock solutions traceable to the National Institute of Standards and Technology (NIST) and ultrapure acids. Instrument drift and matrix effects were monitored and corrected using internal standard elements (^45^Sc, ^89^Y, ^115^In, ^72^Ge, ^103^Rh for ICP-MS, and Y for ICP-AES), which were automatically added from a multi-element mix in 2% HNO3 to each sample and standard prior to analysis.

### 2.6. Calculation of the contribution to recommended nutrient intake

We calculated the contribution of the ten SDF samples to the RNIs for three population groups most vulnerable to micronutrient deficiencies: infants (6–11 months), young children (1–2 years), and women of childbearing age (15–49 years), based on standardized daily intakes. We focused on 11 micronutrients essential for the development and health of these groups and for which reference intake values were available. RNI values for each element and population group were obtained primarily from WHO and FAO (2004). For elements not listed in WHO and FAO (2004), values were retrieved from the (National Academies of Sciences and Medicine, 2019), EFSA (2009, 2012a, 2017) (**Supplementary Table 1)**. When RNIs were unavailable, Adequate Intake (AI) values were used. For iron and zinc, we applied RNI values corresponding to moderate bioavailability, 10% for iron and 30% for zinc, assuming that a diet including fish can be considered a mixed diet (WHO and FAO, 2004). The standardized daily portions of SDF used to calculate contributions were set at 5 g for infants and 10 g for young children and women of childbearing age. These portion sizes were chosen as conservative yet nutritionally relevant estimates, reflecting realistic amounts of dried fish that may be consumed in Madagascar. We evaluated whether the ten SDF samples met FAO/WHO (2001) thresholds for being considered a ‘source of’ or ‘rich in’ micronutrients. A type was classified as a ‘source’ of a micronutrient if a standardized daily portion provided at least 15% of its RNI, and as ‘rich’ if it contributed 30% or more. We also conducted a sensitivity analysis, in which the daily portion of each SDF type ranged from 1 to 100 g for each population group (Ryckman et al. 2021), to identify which daily portion would contribute to 100% of the RNI. Finally, we assessed each SDF sample’s ability to meet RNIs for each population group across all analyzed nutrients by calculating the Mean Adequacy Ratio (MAR), following the methodology described by Hatløy et al. (1998).

### 2.7. Assessment of the risk of exposure to heavy metals

We measured the concentration of four heavy metals: As, Hg, Cd, and Pb. While As and Hg were analyzed in their total forms, only their most toxic forms, inorganic arsenic (iAs) and methylmercury (MeHg), were considered for the risk assessment. iAs represents a higher fraction of total As in freshwater fish (up to 30%) (Saipan et al., 2012) compared to marine fish (0.29–6.34 %) (Bentley and Soebandrio, 2017). As a conservative assumption, we considered that 30% of total As in freshwater fish was present as iAs and 6% in marine fish. Similarly, for Hg we assumed that 100% of total Hg was present as MeHg, as reported proportions range from 11 to 100% (Joiris et al., 1995; Storelli et al., 2003). The risk of heavy metal exposure from consuming SDF was assessed by comparing average concentrations with established tolerable daily intake levels (TDIL) for Cd (0.36 µg/kg body weight [BW]/day; EFSA, 2012a) and MeHg (0.19 µg/kg BW per day; EFSA, 2012b). For iAs and Pb, no safe tolerable intake levels have been established. Therefore, the margin of exposure (MOE) approach was applied using the benchmark dose lower confidence limit (BMDL), which represents the lower bound of the confidence interval for a dose associated with a small increase in the probability of adverse health effects. A BMDL05 value of 0.06 µg/kg BW/day for iAs (EFSA et al., 2024) was applied, corresponding to a 0.5% increase in the risk of adverse health effects. For Pb, the BMDL01 of 0.5 µg/kg BW/day, associated with developmental neurotoxicity in children (EFSA, 2010), was used.

Heavy metal intakes were estimated using average body weight for each population group. As a precautionary measure, and to obtain conservative intake estimates, we used for infants (6–11 months) and children (1–2 years) the highest body weight values reported for boys, *i.e.*, 10.2 kg and 14.6 kg, respectively (WHO, 2007). For women of childbearing age, we used the average body weight of adult women (20–49 years) in sub-Saharan Africa, estimated at 56.5 kg (Garenne, 2011), rounded to 60 kg for calculation purposes, as a reference. The same standardized portions of SDF used for the nutrient intake assessment (Section 2.6) were applied in these estimations.

### 2.8. Statistical analyses

Element concentrations, their contributions to RNIs, and the risk of exposure to heavy metals were compared across the ten samples using a one-way ANOVA followed by post-hoc Tukey tests. Statistical analyses were performed in R version 4.2.1 (R Core Team, 2022).

## 3. Results

### 3.1. Appreciation and consumption of small dried fish

Overall, SDF were widely appreciated, although preferences varied among types. Eight of the nine types had appreciation rates above 50%. Freshwater fish, either sun-dried *Pirina* (F1) or smoked *Ambotsika* (F2), were the most popular, with appreciation rates of at least 83%, followed by pelagic fish, which averaged 62% across the six samples (**Fig. 2**). In contrast, the mixed coastal type *Kalatambo* (M1 and M2) received a very low appreciation score, with only 14% of all respondents reporting that they appreciated it.

**Fig. 2.**
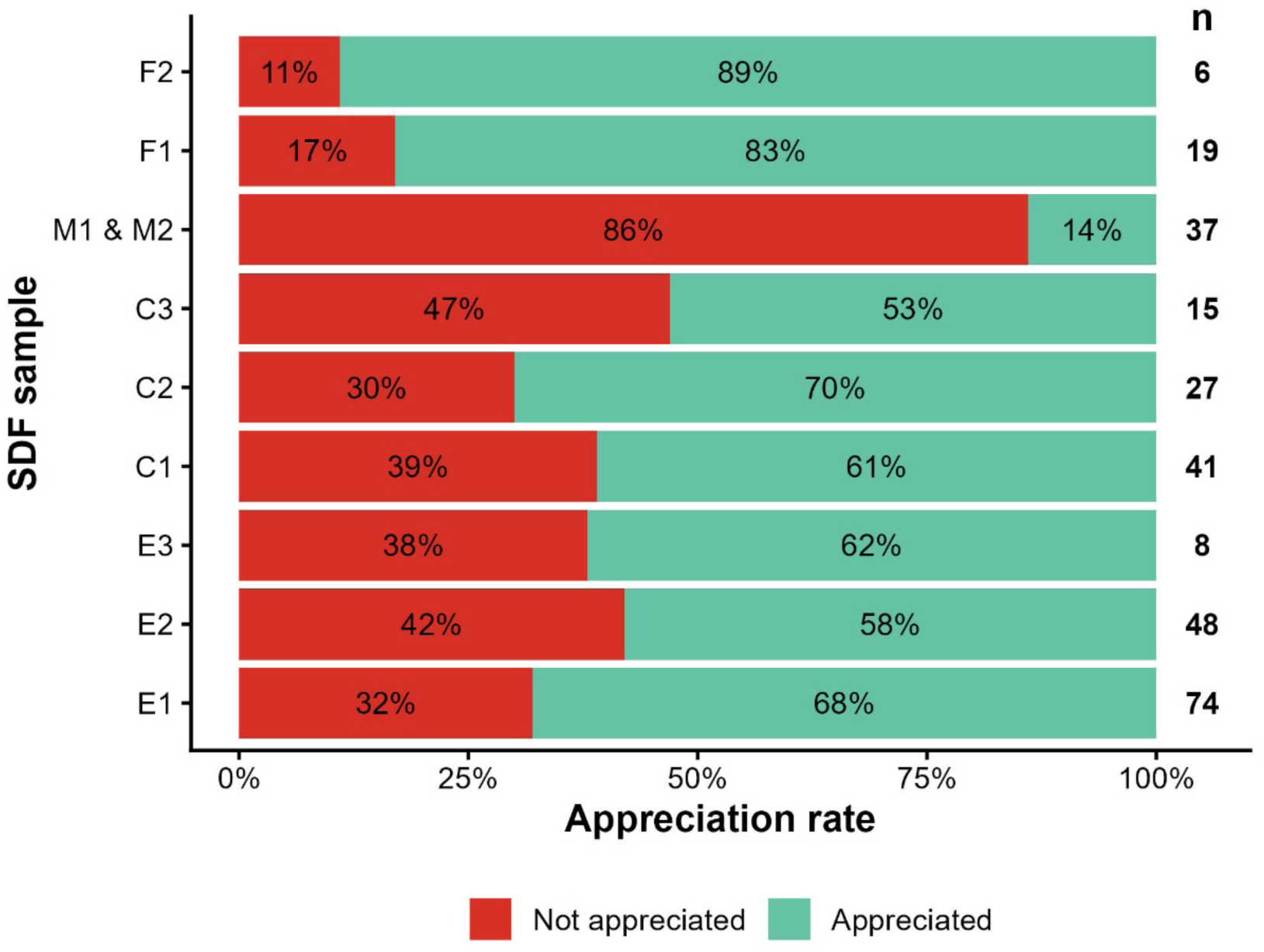
Appreciation rates for the nine types of small dried fish (SDF) considered in the survey. Numbers on the right side (**n**) indicate the total number of respondents per type

SDF were rarely eaten alone and were most often paired with other foods. Six side dishes were identified (**Fig. 3a**), and their use varied according to SDF type. SDF were most commonly consumed with rice, as mentioned by 44% of respondents for *Matsiroky mena hariva* (E3) and by 100% for *Varilava mavokely* (C3). Cassava and corn were also common side dishes, cited by 36% and 22% of respondents, particularly with *Kalatambo* (M1 and M2) and *Matsiroky mena harina* (E3), respectively. Bananas, breadfruit, and sweet potatoes were mentioned less frequently, accounting for only 4% of responses. Regarding preparation methods, SDF were most commonly stewed (42%), fried (27%), or mixed with green vegetables as condiments (23%), while their use in powdered form in sauce was limited (**Fig. 3b**).

**Fig. 3.**
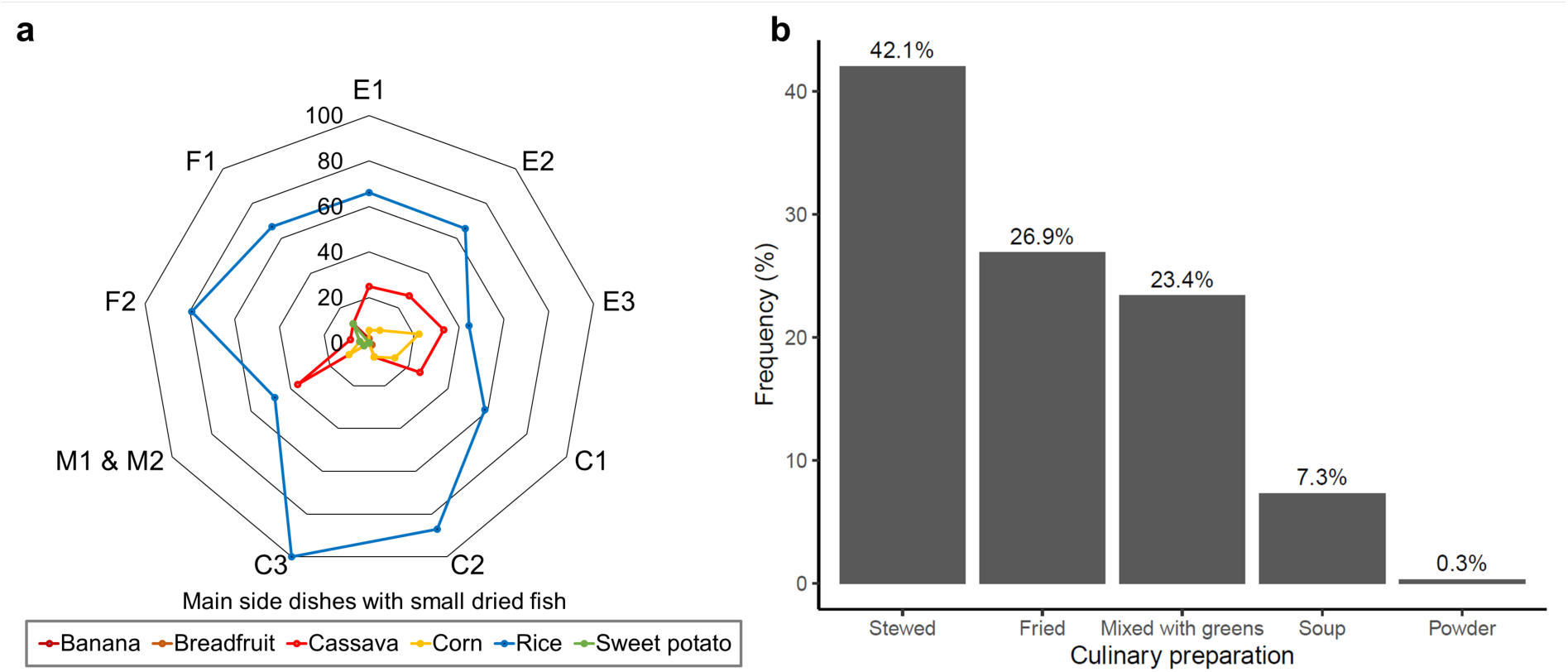
Consumption habits of small dried fish (SDF) in Madagascar. (**a**) Main side dishes eaten with each type of SDF (percentage of respondents). (**b**) Main forms of culinary preparation involving SDF

### 3.2. Taxonomic composition, abundance, and size distribution of small dried fish

The ten SDF samples encompassed a high diversity of fish taxa. Seven samples were dominated by a single species, accounting for 72–100% of the total biomass (**Table 2**). These included two samples dominated by Engraulidae, *Encrasicholina punctifer* (E1) and *Engraulis capensis* (E2), and three dominated by Clupeidae: *Spratelloides gracilis* (C1) and two distinct, yet unidentified *Sauvagella* species (C2 and C3). The freshwater samples included *Gambusia holbrooki* (F1) and an unidentified species from the Eleotridae family (F2). In contrast, samples E3, M1, and M2 displayed higher taxonomic diversity and comprised mixtures of several species (Table 2). Sample E3 consisted of three small pelagic species: *Stolephorus* sp., *Encrasicholina pseudoheteroloba*, and *Sauvagella madagascariensis*. Samples M1 and M2 contained a wide diversity of coastal fish species which, due to identification challenges, were classified at the family level. Sample M1 comprised 12 families, with Gobiidae (19.3%), Labridae (15.2%), and Congridae (10.1%) being the most abundant. By contrast, sample M2 was overwhelmingly dominated by Gobiidae, which accounted for 84.9% of the total biomass. Overall, the ten samples contained 32 distinct fish families. Excluding the mixed coastal fish samples (M1 and M2), we identified 8 families, 13 genera, and 16 putative morphospecies (**Supplementary table 4**).

All fish had SL ranging from 1.1 cm (*Gambusia holbroki*) to 9.2 cm (Congridae) (**Table 2**). Pelagic species had average SL values ranging from 2.6 cm (*Sauvagella* sp. 2) to 5.6 cm (*Engraulis capensis*), while freshwater fish species ranged from 1.5 cm (*Gambusia holbroki* and *Coptodon zillii*) to 3.2 cm (*Eleotris* sp.) (**Table 2**). Coastal fish families also exhibited small average SL values (2.3–9 cm), even in families such as Labridae or Scaridae, which typically include species reaching larger body length.

### 3.3. Micronutrient contents of small dried fish

The average concentrations of the eleven micronutrients varied significantly across the ten samples (**Table 3**). Ca concentrations were high and ranged from 2110 ± 159 (*Matsiroky*, E1) to 6670 ± 920 mg/100 g (*Kalatambo*, M1). All samples had high Se concentrations, ranging from 126 ± 38 (*Pirina*, F1) to 374 ± 39 µg/100 g (*Varilava mavokely*, C3). Fe concentrations were also particularly high, with four samples showing values above 100 mg/100g. Vitamin A was below the detection threshold in all sun-dried samples and was detected only in the smoked sample *Ambotsika* (F2).

**Table 3.**
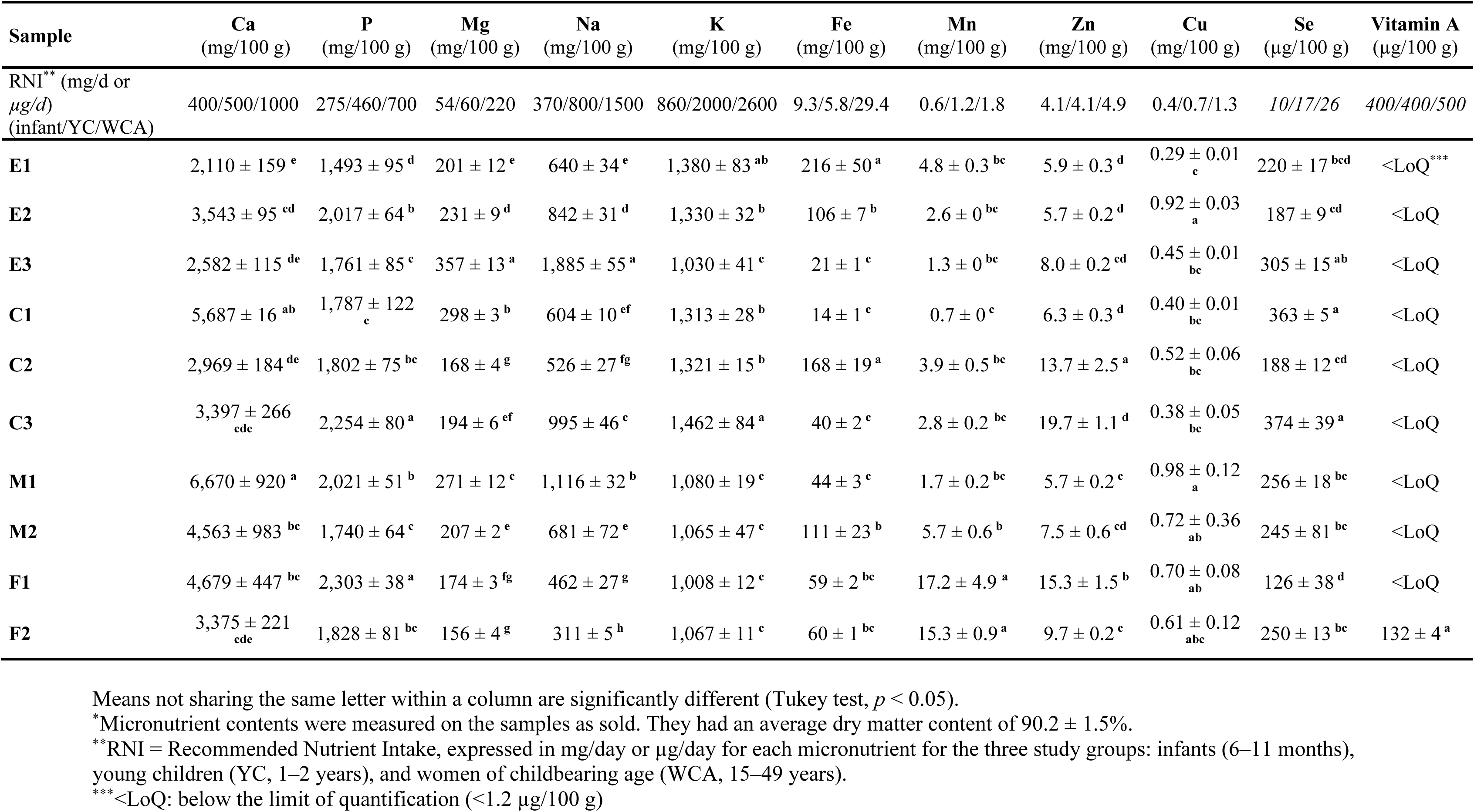
Micronutrient contents* in the ten samples of small dried fish (mean ± standard error)

### 3.4. Heavy metal concentration in small dried fish

Heavy metals were detected in all SDF samples, with concentrations varying markedly across samples (**Table 4**). *Matsiroky* (E1) exhibited the highest concentrations of both As (12.79 ± 0.61 µg/g) and Pb (1.27 ± 0.08 µg/g). *Varilava* (C1) and *Tovy* (E2) showed the highest Cd concentrations, at 1.42 ± 0.01 µg/g and 1.36 ± 0.05 µg/g, respectively. The highest levels of Hg were found in *Ambotsika* (F2) (0.30 ± 0.01 µg/g) and *Varilava mavokely* (C3) (0.28 ± 0.02 µg/g). For the estimated concentration value, inorganic arsenic (iAs) was highest in *Matsiroky* (E1) (0.77 µg/g), followed by *Tovy* (E2) (0.69 µg/g) and Varilava (C1) (0.53 µg/g).

**Table 4.**
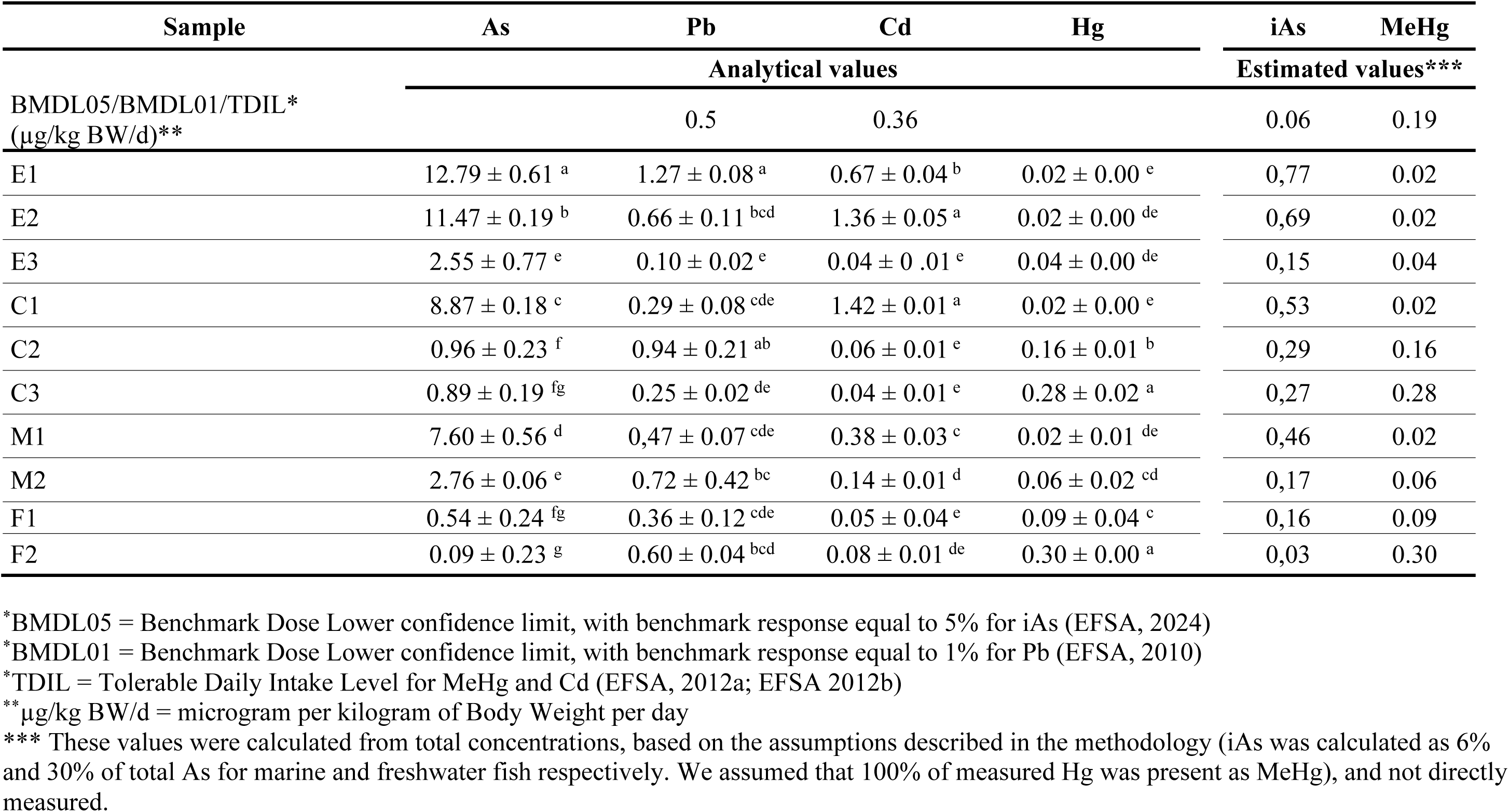
Heavy metal concentrations (µg/g) in the ten samples of small dried fish (mean ± standard error). Means not sharing a letter are significantly different (Tukey test, *P* < 0.05)

### 3.5. Potential contribution of SDF to recommended micronutrient intakes in Madagascar

Despite the relatively small portion sizes considered (5 g for infants and 10 g for children and women), the contributions to RNIs were remarkably high for several micronutrients. A standard portion of SDF qualified as rich in key micronutrients in 124 of the 330 cases considered (nutrient × sample × population group) and as a source of micronutrients in 50 additional cases. Contributions to RNIs varied substantially across nutrients and samples (**Fig. 4**). All samples provided at least 63% of the RNI for Se and 30% of the RNI for Ca in infants and young children, with samples C1 and M1 meeting over 100% of the RNI for both. For women, all SDF samples were rich in Se, and eight samples were rich in Ca. Contributions to the RNI for Fe varied greatly across samples. Nine samples were particularly rich in Fe for young children, with six meeting 100% of their RNI. For infants and women, six and four samples, respectively, were rich in Fe. All SDF samples were either a source of or rich in P for all population groups. In contrast, Mn contributions varied greatly. The two freshwater samples *Pirina* (F1) and *Ambotsika* (F2) met 100% of the RNI in infants and young children and over 85% of the RNI in women. Three other samples, *Mena hariva* (C2)*, Matsiroky* (C1), and *Kalatambo* (M2), were also rich in Mn for infants and children. Seven samples were rich in Mg for young children, but only one (*Matsiroky mena hariva*, E3) for infants. Zn contributions varied substantially across groups and samples (7–48%). Three samples were rich in Zn for young children (33–48%) and two for women (31 and 40%). For infants, three samples, *Mena hariva* (C2)*, Varilava mavokely* (C3), and *Pirina* (F1), were a source of Zn. Contributions to Na, Cu, and K recommended intakes were generally low across all samples and population groups. Only the smoked sample *Ambotsika* (F2) slightly contributed to vitamin A requirements, providing 2–3% of the RNI.

**Fig. 4.**
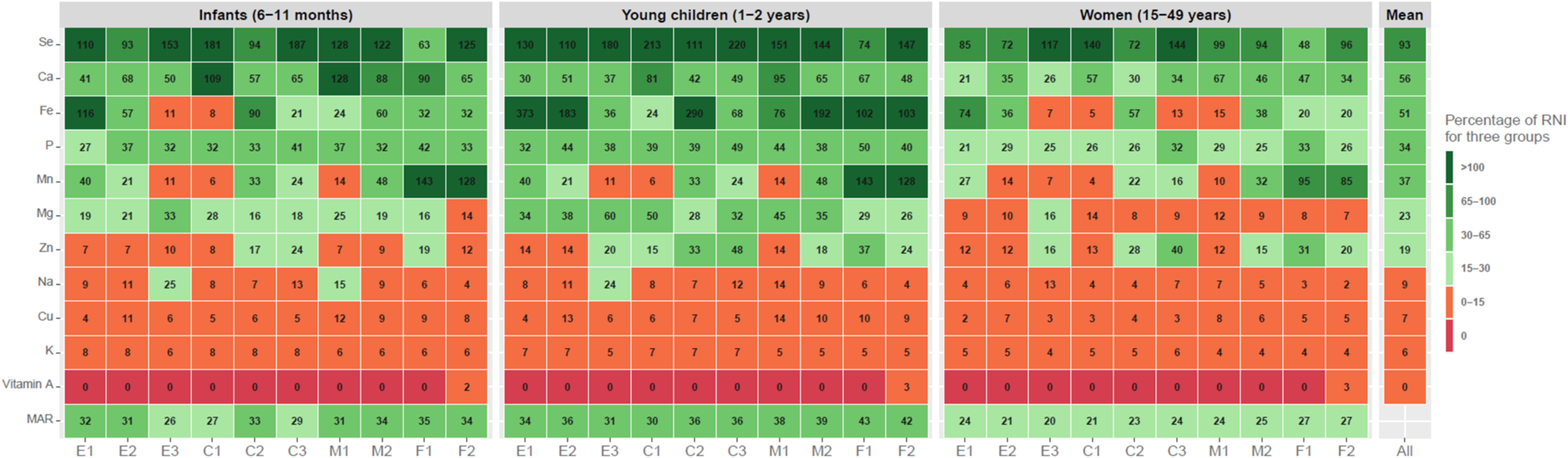
Contribution of a standard portion of small dried fish (5 g for infants; 10 g for young children and women) from 10 samples to the RNI of 11 key micronutrients. Each panel corresponds to one population group: infants (6–11 months), young children (1–2 years) and women of childbearing age (15–49 years). Black numbers indicate the percentage of the RNI. ‘Mean’ corresponds to the average percentage of RNI across all samples and population groups. ‘MAR’ corresponds to the mean adequacy ratio for each sample and population group

**Fig. 5.**
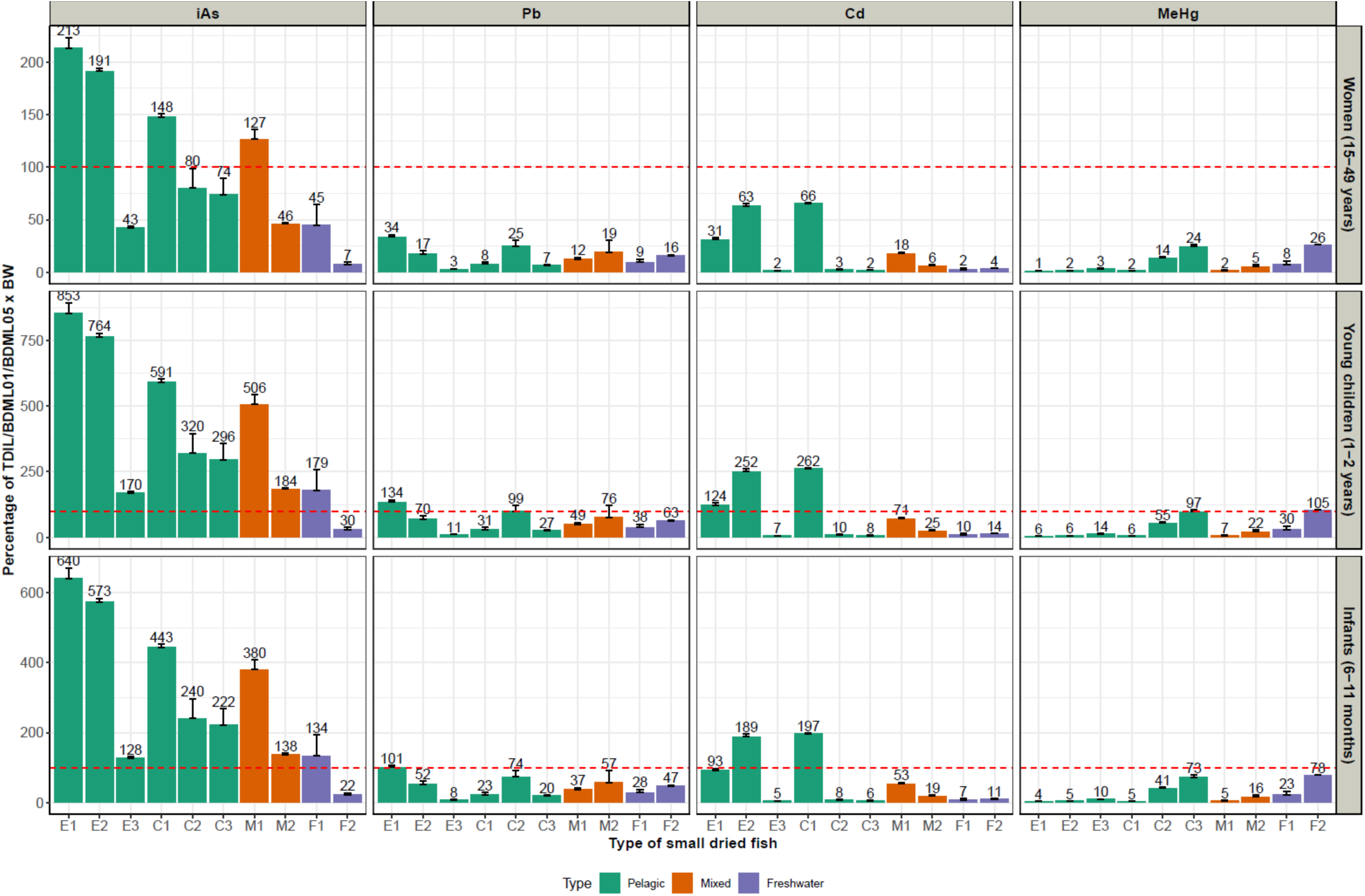
Estimated contribution of a portion of small dried fish to toxicological reference values, expressed as a percentage of TDIL for Cd and MeHg or BMDL values for iAs and Pb, for infants (mean body weight: 10.2 kg), young children (mean body weight: 14.6 kg) and women (mean body weight: 60 kg). The horizontal red dashed line represents 100% of the TDIL or BMDL reference value. Bars with error bars represent estimated means ± standard error. BMDL, benchmark dose lower confidence limit; TDIL, tolerable daily intake level

### 3.6. Heavy metal exposure

The ten SDF samples showed substantial variation in heavy metal concentrations (**Table 4**). Consumption of a standard portion, 5 g for an infant with an average body weight of 10.2 kg, and 10 g for a young child (14.6 kg) or a woman (60 kg), exceeded the reference values for certain samples (**Fig. 6**). Infants and young children showed higher exposure risks than women due to their lower body weight and proportionally higher intake. In general, the estimated iAs intake from standard portions of most samples greatly exceeded the BMDL05 threshold, particularly in infants and young children. Only one freshwater sample, *Ambotsika* (F2), was an exception, with a contribution of less than 30% of the reference value. In contrast, consumption of a standard portion of most SDF type did not exceed the TDIL for estimated MeHg across all groups, except for F2, which reached 105% in young children. Cd TDIL exceedances were also observed, especially among infants and young children consuming pelagic SDF types such as *Tovy* (E2) and *Varilava* (C1). Contributions of Pb remained below 100% overall, suggesting a lower risk for these elements. The freshwater sample F2 posed lower risks for iAs, Pb, and Cd, but higher risks for MeHg.

## 4. Conclusion

Processed aquatic foods, including dried and smoked fish, are increasingly recognized as important foods in LMICs, as they combine affordability, storability, and micronutrient density in ways that make them particularly relevant for addressing food and nutrition insecurity (Byrd et al., 2021; Belton et al., 2022; Robinson et al., 2025). To our knowledge, this study is the first to simultaneously assess the availability, diversity, consumer preferences, nutritional benefits, and potential health risks of SDF in Madagascar. By combining market surveys, taxonomic analyses, micronutrient profiling, and contamination assessment, we show that SDF are widely available across both coastal and inland regions and constitute a key animal-source food for low-income households.

### 4.1. Small dried fish as a widespread aquatic food in Malagasy food systems

The widespread presence of SDF across all surveyed markets confirms their central role in local food systems. Their availability along Madagascar’s main transportation axis (RN7), including inland areas, highlights their potential to overcome geographical constraints that limit access to fresh aquatic food. In addition, their affordability and the possibility of purchasing them in small portions make SDF more accessible than other animal-source foods, such as meat (Dürr and Ratompoarison, 2021), particularly for low-income households (Ndrianaivo et al., 2015).

Our findings reveal a high and largely hidden taxonomic diversity of SDF in Malagasy markets, with 33 distinct fish families identified among the nine most common commercial types, representing 13.4% of Madagascar’s known fish families (Fricke et al., 2018). However, small pelagic families, particularly Engraulidae and Clupeidae, dominated the supply. Consumer preferences further revealed important differences among types. The freshwater types *Pirina* and *Ambotsika* were the most appreciated despite their higher price, suggesting that taste or cultural preferences may outweigh cost considerations. However, affordability remained a key constraint, as some consumers reported choosing less expensive types over preferred but more costly ones. Beyond freshwater species, small pelagic species were also well appreciated, whereas mixed coastal types (*Kalatambo*) were the least appreciated.

### 4.2. Small dried fish as a highly nutrient-dense aquatic food for addressing deficiencies

Despite substantial national fish production, Madagascar shows a marked mismatch between aquatic food availability and consumption, with low per capita intake and persistent micronutrient deficiencies, especially among vulnerable populations (INSTAT and ICF, 2022; ONN, 2023). In this context, SDF may play a critical role, as their affordability, storability, and widespread distribution make them accessible sources of essential micronutrients often lacking in the diets of infants and young children. Our results show that SDF are highly nutrient-dense foods, particularly with respect to Ca, Fe, Se, Mn, and Zn. Even small portions (5 and 10 g) provided substantial contributions to RNIs and often qualified as a source of, or rich in, several key micronutrients for vulnerable groups such as infants, young children, and women of reproductive age. These findings are consistent with previous studies in other LMICs, which have highlighted the nutritional importance of small fish consumed whole (Siddhnath et al., 2022; Wessels et al., 2023; Mamun et al. 2024; Reza et al., 2024). The high micronutrient density observed across SDF is largely explained by the fact that these products are prepared and consumed whole, including bones and viscera, that are rich in minerals. However, our results also reveal significant variability in micronutrient content across species, processing methods, and ecological origins.

The concentrations observed in our samples fell within, and in some cases exceeded, the ranges reported for small fish consumed whole, particularly for iron (Reza et al., 2024; Siddhnath et al., 2022; Sun et al., 2025) and calcium (Mamun et al., 2024; Sun et al., 2025).

The high iron levels observed in some samples may reflect differences in processing and handling conditions, which can influence iron concentrations in dried fish products. In contrast, vitamin A was detected only in the smoked freshwater sample. The absence of vitamin A (retinol) in sun-dried samples likely reflects degradation during drying, as this vitamin is thermolabile and sensitive to ultraviolet radiation (Roos et al., 2002, 2007). Previous studies have reported detectable vitamin A in similar species, such as *Spratelloides gracilis* (Mamun et al., 2024). The fact that vitamin A was detected only in the smoked sample *Ambotsika* further suggests that processing, storage, and handling conditions play a key role in shaping micronutrient composition.

Overall, our results showed substantial variability among SDF types, with some combining particularly high micronutrient densities with strong consumer appreciation. These findings support the potential of SDF as part of food-based strategies to address micronutrient deficiencies in Madagascar, where diets remain dominated by starchy staples and access to diverse animal-source foods is limited.

### 4.3. Variability in contamination risks among small dried fish

Despite their nutritional benefits, SDF also represent a potential source of exposure to environmental contaminants. As, Cd, Hg, and Pb were detected in all samples, with substantial variation among them. In particular, estimated exposure to Cd and iAs exceeded acceptable reference values for several pelagic species at standard portion sizes, especially for infants and young children. Consumption of a standard portion of two pelagic SDF types (*Matsiroky* and *Varilava)* exceeded the Cd TDIL in both infants and young children. Likewise, consumption of standard portions of nine SDF samples led to estimated iAs intakes in infants and young children that exceeded the BMDL05-based reference value by 1.3- to 8.5-fold. The particularly high frequency of iAs exceedances may partly reflect the use of a conservative benchmark value of 0.06 µg/kg body weight/day proposed by EFSA et al. (2024), whereas other studies, such as Wessels et al. (2023), used a higher reference value of 0.3 µg/kg body weight/day. Nonetheless, our findings are consistent with previous studies reporting heavy metal contamination in dried fish products from other regions (Akter et al., 2019; Anisha et al., 2023; Mamun et al., 2024; Sun et al., 2025), although some of the concentrations observed in our samples were higher. High iAs concentrations may reflect environmental contamination in fishing areas or bioaccumulation within marine food webs (Jordão et al., 2002). In Madagascar, mining and other extractive activities have been associated with pollution of water, soils, and air, as well as increased erosion and sedimentation of waterways, although the extent of chemical contamination remains poorly documented at the national scale (GAHP, 2018; Devenish et al., 2023; Zaehringer et al., 2024). In our case, six of the samples analyzed were harvested near Saint Augustin, where the seasonal Onilahy river, one of the largest rivers in south-west Madagascar, flows into the ocean. This river can transport substantial quantities of sediment, which could contribute to contaminant transport and accumulation in nearby marine ecosystems. However, contamination risks were not uniform across SDF types. Freshwater species and some pelagic species showed comparatively lower exposure risks, highlighting the importance of species-level differentiation. This variability underscores the need to move beyond general assumptions about the safety of dried fish and to better understand the ecological and environmental drivers of contamination.

### 4.4. The need to balance nutritional benefits and food safety risks

Our results highlight the trade-offs between nutritional benefits and contaminant exposure across SDF types. The most abundant and widely appreciated pelagic SDF (*Matsiroky*, *Tovy* and *Varilava*) provided high micronutrient densities but also showed the greatest exceedances of acceptable upper limits for iAs or Cd. In contrast, some less abundant or more expensive types (*Pirina*, *Ambotsika*, and *Mena hariva*) combined strong nutritional profiles with comparatively lower contamination risks. This suggests that promoting SDF consumption to address micronutrient deficiencies requires a more nuanced, integrated, and type-specific approach. For vulnerable groups such as infants and young children, both portion size and frequency of consumption should be carefully considered in order to balance nutritional benefits against potential risks. These findings also highlight the need for more integrated studies that jointly assess nutritional quality and food safety.

## 5. Conclusion

SDF represent a promising aquatic food for improving access to essential micronutrients in LMICs. In Madagascar, they are are widespread, affordable, and can be stored for long periods, making them particularly relevant for rural and inland populations with limited access to fresh animal-source foods. Yet, maximizing their nutritional potential requires targeted interventions to limit heavy metal exposure. These may include promoting safer SDF types, improving post-harvest handling and processing practices to reduce contamination, and encouraging the use of powdered fish to facilitate consumption by young children. Raising awareness among consumers about both the benefits and risks associated with different SDF types will also be essential.

## Authors’ bios

### Léono D. Todimazava

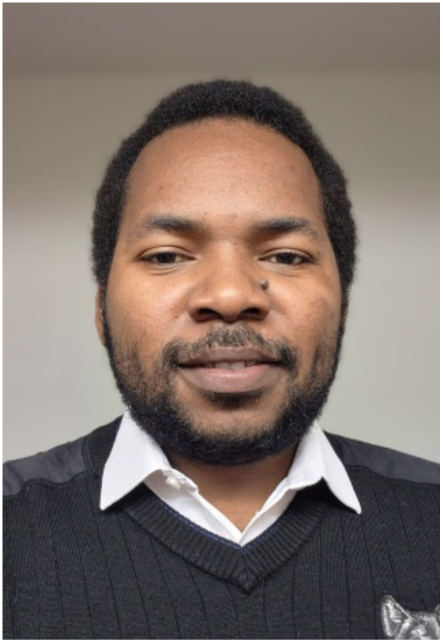

Léono D. Todimazava is a young searcher in marine and fisheries sciences, specializing in the sustainable use of aquatic resources. He holds a Master’s degree from the Institut Halieutique et des Sciences Marines (IH.SM) at the University of Toliara and is currently pursuing a PhD study which aims to assess the contribution of aquatic products to food and nutritional security, particularly among vulnerable populations. Since his Master’s degree, his research has taken an integrated approach combining fisheries, biodiversity and nutrition, with a focus on fisheries productivity, resource accessibility and the nutritional quality of aquatics products (both fresh and processed), in order to identify sustainable ways to improve food and nutritional security.

### Maria J. Darias

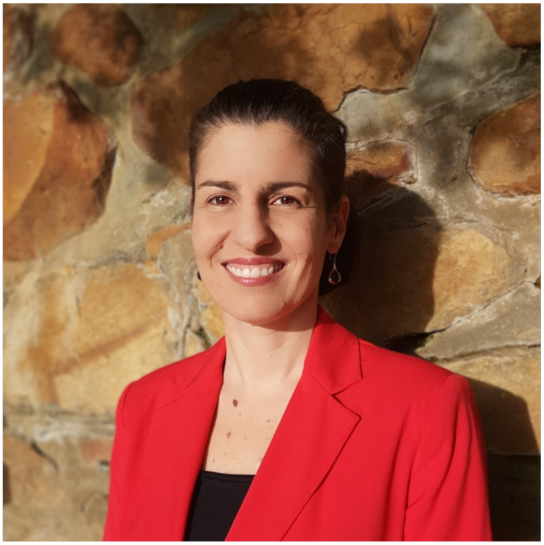

Maria J. Darias is a tenured Research Professor at the French National Research Institute for Sustainable Development (IRD), where she pursues inter- and transdisciplinary research to advance sustainable and nutrition-sensitive aquaculture and aquatic food systems in the Global South. Her research links aquatic animal nutrition and physiology with aquatic food composition and safety, with the aim of improving the nutritional value and sustainability of farmed aquatic foods. She holds a PhD in Animal Biology from the University of Cádiz, Spain, and a Habilitation (HDR) from the University of Montpellier, France. She coordinates AfriMAQUA, a nutrition-sensitive marine aquaculture network in Africa, and co-leads LIMAQUA, a South African–French partnership program for interdisciplinary research and capacity building addressing nutritional and sustainability challenges in marine aquaculture.

### Claire Mouquet-Rivier

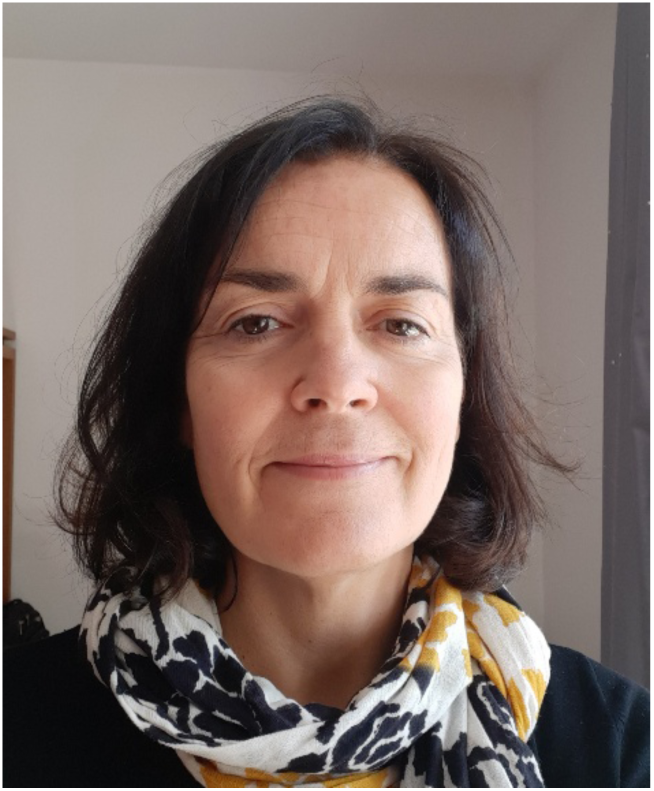

Claire Mouquet-Rivier, PhD in food science and nutrition, works at the French National Research Institute for Sustainable Development (IRD) as a senior researcher in public health nutrition. She heads the ‘Food, Nutrition and Health’ team within the QUALISUD joint research unit. Her work focuses on designing and evaluating integrated strategies based on locally available foods to prevent undernutrition and micronutrient deficiencies. She is particularly interested in the design of complementary foods suitable for young children: increasing energy density as well as the content and bioavailability of micronutrients. She carries out this work in collaboration with institutional partners in sub-Saharan Africa through field studies characterising the dietary and nutritional intakes of vulnerable groups. She is co-author of 80 publications in peer-reviewed international journals.

### Jamal Mahafina

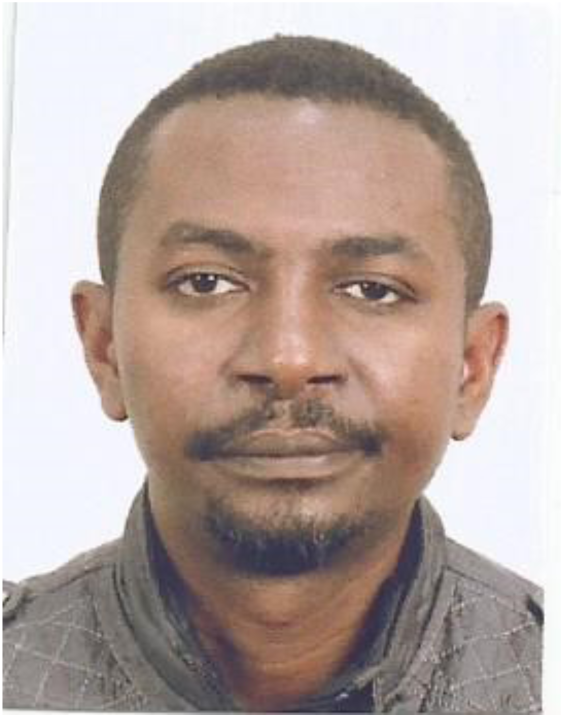

Jamal Mahafina is a Professor at the Fisheries and Marine Sciences Institute at the University of Toliara and head of the Fisheries and Aquatic Resources Management Laboratory within the institute. For more than 15 years, he has worked in partnership on research topics related to small-scale fisheries and reef fish in Madagascar to promote both the sustainable management of marine resources and food and nutritional security. His work addresses different life stages of reef fish (post-larvae, juveniles, adults).

### Thomas Lamy

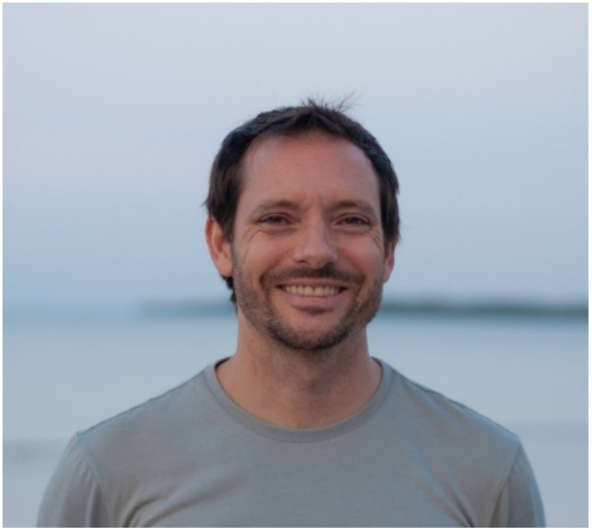

Thomas Lamy is a marine ecologist at the French Research Institute for Sustainable Development (IRD). His research focuses on the biodiversity, resilience, and conservation of coastal ecosystems, with particular attention to the role of aquatic foods in nutrition and food security. He combines ecological, genetic, and social approaches to understand how marine ecosystems respond to global change and support food security and the livelihoods of coastal communities.

## Tables

### Statements and Declarations

The authors have no competing interests to declare that are relevant to the content of this article.

## Data Availability

All data produced are available in Zenodo: 10.5281/zenodo.22115072

https://zenodo.org/records/22115072

## Acknowledgments

We are very grateful to Dominique Ponton for inspiring this study and for providing the computer and camera that made it possible. We sincerely thank Johanès Tsilavonarivo and Professor Anusha Rajkaran for transporting the samples to South Africa. We also thank Jean-Dominique Durand for his advice on DNA barcoding and for blasting our sequences against the 12S MADFI reference database that he developed. We are grateful to Tsipy Romano, Duphérino, and Rasoanirina Marie Francéline for their assistance with the laboratory work. This work was publicly funded through ANR (the French National Research Agency) under the “Investissements d’avenir” program with the reference ANR-16-IDEX-0006 (Project FISHTAIL), as well as by the IRD LMI (joint international laboratory) MIKAROKA.

## Ethics approval

Data collection was conducted in accordance with the advisory opinion of the Research Ethics Committee at the University of Montpellier (UM 2023-040bis). This research was conducted under research permit 181/22/MEDD/SG/DGGE/DAPRNE/SCBE.Re, issued by the Direction des Aires Protégées, des Ressources Naturelles Renouvelables et des Ecosystèmes in Madagascar.

