## Supplementary information for "Small dried fish as an affordable source of key micronutrients in Madagascar: nutritional benefits and contamination risks"

**Supplementary Table 1** Summary of the main micronutrients considered in this study. For each nutrient, the Recommended Nutrient Intake (RNI) or Adequate Intake (AI), together with the corresponding reference, is provided for three population groups: infants (6–11 months), young children (1–2 years) and women of reproductive age (15–49 years). When available, the extent of deficiency for each nutrient in Madagascar is also indicated. The importance of each nutrient for human health is briefly described

| **Symbol** | **Unit** | Infants (6–11 months) | Young children (1–2 years) | Women (15–49 years) | **Reference for RNI/AI** | **Deficiency in Madagascar** | **Importance for human health** | **References for health importance** |
|---|---|---|---|---|---|---|---|---|
| | | | **RNI or AI*** | | | | | |
| **Na** | mg/d | 370 | 800 | 1,500* | National Academies of Sciences and Medicine (2019) | NA | Maintains extracellular volume and plasma osmolality. Is also an essential determinant of cell membrane potential and the active transport of molecules across cell membranes. | Lang et al. (1998); Shah and Mandiga (2024) |
| **Mg** | mg/d | 54 | 60 | 220 | WHO and FAO (2004) | High (Passarelli et al., 2024) | Essential for the proper activity of many biochemical and physiological processes, including DNA replication, transcription and translation. As a coenzyme or an activator for many enzymes involved in energy metabolism, protein synthesis and | Farias et al. (2020); Fiorentini et al. (2021); Glasdam et al. (2016) |

| | | | | | | | | |
|---|---|---|---|---|---|---|---|---|
| | | | | | | | maintenance of the electrical potential of nervous tissues and cell membranes. As such, it is important for proper cerebral function. | |
| **P** | mg/d | 275 | 460 | 700 | Institute of Medicine (2000) | NA | Plays an essential role in skeletal development, mineral metabolism, and various cellular functions, including intermediary metabolism and energy transfer mechanisms. | Serna and Bergwitz (2020); Farag et al. (2023) |
| **K** | mg/d | 700 | 2,000 | 2,600* | Institute of Medicine (2000) | NA | Plays a key role in maintaining cell function. Contributes to osmolarity and plays a major role in the distribution of fluids inside and outside cells. Plays a role in regulating water balance and acid–base balance in blood and tissues. | Palmer (2015); Udensi and Tchounwou (2017) |
| **Ca** | mg/d | 260 | 700 | 1,300 | National Academies of Sciences and Medicine (2019) | Potential deficiency (Unicef, 2023) | Has a structural role and is needed for tissue rigidity, strength and elasticity. Participates in whole-body mineral homeostasis through the processes of bone formation and resorption. | Farag et al. (2023); Farias et al. (2020) |
| **Fe** | mg/d | 9.3 | 5.8 | 31 | WHO and FAO (2004) | High (Unicef, 2023) | Serves as a carrier of oxygen to the tissues by red blood cells (hemoglobin), as a transport medium for electrons within cells, and as an integrated part of important enzyme systems in various tissues. Is crucial for normal growth and development, particularly in infants, and for maintaining immune function. | Abbaspour et al. (2014; Farias et al. (2020) |
| **Zn** | mg/d | 4.1 | 4.1 | 7.2 | WHO and FAO (2004) | Potential deficiency (Unicef, 2023) | Involved in many biochemical and physiological processes linked to brain growth and function, as well as cellular metabolism. Structural stabilizer, fatty | Farias et al. (2020); Sheffler et al. (2024) |

| | | | | | | | | |
|---|---|---|---|---|---|---|---|---|
| | | | | | | | acid metabolism and antioxidant defense. | |
| **Se** | μg/d | 10 | 17 | 26 | WHO and FAO (2004) | High (Passarelli et al., 2024) | Has antioxidant activity and plays a role in oxidation-reduction reactions and thyroid hormone metabolism. | Rayman (2000); Zhang et al. (2023) |
| **Cu** | mg/d | 0.4 | 0.7 | 1.3 | EFSA (2017) | NA | Contains the key enzyme (a cuproenzyme) involved in the cross-linking of connective tissues, such as collagen and elastin fibers, which are structural components present in tissues such as skin, bone and blood vessels. It is essential for the mineralization of the skeleton and plays a role in regulating glucose metabolism. | Rucker et al. (1998); Upadhaya and Kim (2020) |
| **Mn** | mg/d | 0.6 | 1.2 | 1.8 | WHO and FAO (2004) | NA | Required for proper immune function, regulation of blood sugar and cellular energy, bone growth, blood coagulation and hemostasis, and defense against reactive oxygen species. It is a component of metalloenzymes and is involved in oxidation-reduction reactions and proteoglycan synthesis. | Horning et al. (2015); Li and Yang (2018); Thirunavukarasu et al. (2022) |
| **Vitamin A** | μg/d (RE) | 400 | 400 | 600 | WHO and FAO (2004) | Potential deficiency (Unicef, 2023) | Essential for maintaining the proper functioning of the visual system. Promotes growth, development and maintenance of epithelial cell integrity, and supports immune and reproductive function. | Huang et al. (2018); Thirunavukarasu et al. (2022) |

*AI: Adequate intake

RE: Retinol Equivalent

**Supplementary Table 2** Overview of the market survey of small dried fish (SDF) conducted in different cities along National Road 7 (NR7), from coastal Toliara to inland Antananarivo

| City | Market | Market day | Number of vendors present | Number of vendors surveyed | Number of SDF batches recorded | Number of SDF batches sampled |
|---|---|---|---|---|---|---|
| **Toliara** | Scama | Daily | 6 | 6 | 14 | 5 |
| | Bazar Be | Daily | 1 | 1 | 1 | 1 |
| **Andranovory** | Bazar Andranovory | Sunday | 8 | 5 | 8 | 5 |
| **Sakaraha** | Bazar Be | Daily | 5 | 5 | 22 | 12 |
| | Tsenavao | Saturday[a] | 31 | 9 | 25 | 12 |
| **Ilakaka** | Magarahara | Daily | 6 | 6 | 18 | 9 |
| | Stationnement | Daily | 4 | 4 | 4 | 3 |
| **Ranohira** | Bazar Ranohira | Friday[a] | 8 | 8 | 25 | 10 |
| **Andiolava** | Bazar Andiolava | Saturday | 5 | 5 | 20 | 0 |
| **Ihosy** | Bazar Tanambao | Monday[a] | 13 | 9 | 28 | 12 |
| **Ambalavao** | Antanimalalaka | Wednesday[a] | 21 | 7 | 16 | 7 |
| **Fianarantsoa** | Anjoma | Friday[a] | 29 | 8 | 46 | 11 |
| **Ambositra** | Iajaky | Saturday[a] | 11 | 11 | 19 | 7 |
| **Antsirabe** | Asabotsy | Saturday[a] | 35[b] | 12 | 50 | 10 |
| **Antananarivo** | Tsenabe Isotry | Friday[a] | 92 | 9 | 46 | 14 |
| | Andravoahangy | Wednesday[a] | 38 | 7 | 47 | 13 |
| **TOTAL** | | | **313** | **112** | **389** | **131** |

[a]This represents the main market day, characterized by a large number of SDF sellers and consumers from other towns or villages. However, this does not exclude sales on other days.

[b]This market was considerably larger than the others. Due to time constraints, the number given is an underestimate.

**Supplementary Table 3** Diversity of of small dried fish batches observed for each vendor in the different markets surveyed along National Road 7. A unique code was assigned to each vendor in order to preserve anonymity. For each batch observed, the processing method was recorded and the corresponding Malagasy name was noted, reviewed, and harmonized by consensus according to batch composition

| City | Market | Vendor code | Processing method | Malagasy name |
|---|---|---|---|---|
| Toliara | Scama | V1 | Sun drying | Tovy |
| | | | | Tovy |
| | | | | Bemaso |
| | | V2 | Sun drying | Matsiroky |
| | | | | Matsiroky |
| | | | | Tovy |
| | | V3 | Sun drying | Tovy |
| | | | | Kabiliky |
| | | V4 | Sun drying | Matsiroky |
| | | | | Kabiliky |
| | | V5 | Sun drying | Tovy |
| | | V6 | Sun drying | Bemaso |
| | | | | Tovy |
| | | | | Matsiroky |
| | Bazar Be | V7 | Sun drying | Matsiroky |
| Andranovory | Bazar Andranovory | V8 | Sun drying | Kalatambo |
| | | | | Kabiliky |
| | | | | Kabiliky |
| | | V9 | Sun drying | Tovy |
| | | V10 | Sun drying | Varilava |
| | | V11 | Sun drying | Tovy |
| | | V12 | Sun drying | Tovy |
| | | | | Tovy |
| Sakaraha | Bazar Be | V13 | Sun drying | Kabiliky |
| | | | | Kalatambo |
| | | | | Kalatambo |
| | | | | Matsiroky |
| | | | | Tovy |
| | | | | Tovy |
| | | V14 | Sun drying | Matsiroky |
| | | | | Matsiroky |
| | | V15 | Sun drying | Viandrano |
| | | | | Kalatambo |
| | | | | Matsiroky |
| | | V16 | Sun drying | Tovy |
| | | | | Matsiroky |

| | | | | |
|---|---|---|---|---|
| | | | | Matsiroky |
| | | | | Matsiroky |
| | | | | Kabiliky |
| | | | | Kalatambo |
| | | | | Viandrano |
| | | | | Kalatambo |
| | | | | Varilava |
| | | V17 | Sun drying | Kalatambo |
| | | | | Tovy |
| | Tsenavao | V18 | Sun drying | Haroky |
| | | | | Varilava |
| | | V19 | Sun drying | Kalatambo |
| | | | | Bemaso |
| | | | | Kalatambo |
| | | V20 | Sun drying | Bemaso |
| | | | | Kalatambo |
| | | | | Lanora |
| | | | | Fitse |
| | | V21 | Sun drying | Kalatambo |
| | | | | Matsiroky |
| | | | | Tovy |
| | | V22 | Sun drying | Matsiroky |
| | | V23 | Sun drying | Matsiroky |
| | | | | Matsiroky |
| | | | | Kalatambo |
| | | | | Tovy |
| | | V24 | Sun drying | Tovy |
| | | | | Kabiliky |
| | | V25 | Sun drying | Antseradava |
| | | | | Tovy |
| | | | | Varilava |
| | | V26 | Sun drying | Tovy |
| | | | | Varilava |
| | | | | Matsiroky |
| Ilakaka | Mangarahara | V27 | Sun drying | Lily |
| | | | | Kalatambo |
| | | | | Matsiroky mena hariva |
| | | | | Matsiroky |
| | | | | Kalatambo |
| | | V28 | Sun drying | Matsiroky mena hariva |
| | | | Smoking | Tilapia |

| | | | | |
|---|---|---|---|---|
| | | V29 | Sun drying | Matsiroky mena hariva |
| | | V30 | Sun drying | Tovy |
| | | | | Kabiliky |
| | | | | Tovy |
| | | | | Tovy |
| | | | | Tovy |
| | | | | Matsiroky mena hariva |
| | | V31 | Sun drying | Matsiroky |
| | | | | Tilapia |
| | | V32 | Sun drying | Tovy |
| | | | | Varilava |
| | Stationnement | V33 | Sun drying | Matsiroky |
| | | V34 | | Kalatambo |
| | | V35 | | Matsiroky |
| | | V36 | | Kalatambo |
| Ranohira | Bazar Ranohira | V37 | Sun drying | Matsiroky mena hariva |
| | | | | Kabiliky |
| | | | | Tovy |
| | | | | Matsiroky |
| | | | | Varilava |
| | | V38 | Smoking | Varilava |
| | | | | Matsiroky |
| | | | | Kalatambo |
| | | V39 | Smoking | Kalatambo |
| | | V40 | Smoking | Varilava |
| | | | | Bemena |
| | | | | Ambila |
| | | | | Matsiroky mena hariva |
| | | | | Varilava |
| | | | | Mena hariva |
| | | | | Kabiliky |
| | | | | Varilava |
| | | | | Kabiliky |
| | | V41 | Smoking | Vily |
| | | V42 | Sun drying | Kalatambo |
| | | | | Kalatambo |
| | | V43 | Sun drying | Matsiroky |
| | | | | Kalatambo |
| | | V44 | Sun drying | Kalatambo |
| | | | | Kalatambo |
| Andiolava | Bazar Andiolava | V37 | Sun drying | Matsiroky mena hariva |
| | | | | Kabiliky |
| | | | | Tovy |

| | | | | |
|---|---|---|---|---|
| | | | | Matsiroky |
| | | | | Varilava |
| | | V38 | Sun drying | Varilava |
| | | | | Matsiroky |
| | | | | Kalatambo |
| | | V40 | Sun drying | Varilava |
| | | | | Bemena |
| | | | | Ambila |
| | | | | Matsiroky mena hariva |
| | | | | Varilava |
| | | | | Mena hariva |
| | | | | Kabiliky |
| | | | | Varilava |
| | | | | Kabiliky |
| | | V41 | Smoking | Vily |
| | | V43 | Sun drying | Matsiroky |
| | | | | Kalatambo |
| Ihosy | Tanambao | V45 | Smoking | Ambotsika |
| | | | | Pirina |
| | | | Sun drying | Pirina |
| | | V46 | Smoking | Fiamainty |
| | | V47 | Sun drying | Matsiroky |
| | | | | Kalatambo |
| | | V48 | Smoking | Ambotsika |
| | | | Sun drying | Matsiroky |
| | | | | Kalatambo |
| | | | | Kalatambo |
| | | | | Vily |
| | | | Smoking | Ambotsika |
| | | V49 | Sun drying | Kalatambo |
| | | | | Mena hariva |
| | | | | Matsiroky |
| | | | | Ambotsika |
| | | V50 | Smoking | Ambotsika |
| | | | Sun drying | Kalatambo |
| | | | | Tovy |
| | | | | Kalatambo |
| | | | | Matsiroky |
| | | V51 | Sun drying | Karara |
| | | | | Kabiliky |
| | | | | Tovy |
| | | V52 | Sun drying | Kalatambo |
| | | V53 | Smoking | Pirina |

| | | | | |
|---|---|---|---|---|
| | | | | Ambotsika |
| | | | Sun drying | Kalatambo |
| Ambalavao | Antanimalalaka | V54 | Sun drying | Kalatambo |
| | | | | Matsiroky |
| | | V55 | Sun drying | Matsiroky |
| | | | | Kabiliky |
| | | | | Kalatambo |
| | | V56 | Sun drying | Kalatambo |
| | | | | Matsiroky |
| | | V57 | Sun drying | Matsiroky |
| | | | | Karara |
| | | V58 | Sun drying | Kabiliky |
| | | | | Varilava |
| | | | | Matsiroky |
| | | | | Matsiroky |
| | | | | Kalatambo |
| | | V59 | Sun drying | Pirina |
| | | V60 | Smoking | Pirina |
| Fianarantsoa | Anjoma | V61 | Sun drying | Karara |
| | | | | Amba amba |
| | | | | Gogo |
| | | | | Lily |
| | | V62 | Sun drying | Varilava |
| | | | | Matsiroky |
| | | | | Amba amba |
| | | | | Karara |
| | | V63 | Sun drying | Amba amba |
| | | | | Karara |
| | | | | Varilava |
| | | | Fermented | Lily |
| | | | | Gogo |
| | | V64 | Sun drying | Kabiliky |
| | | | | Karara |
| | | | | Matsiroky |
| | | | | Varilava |
| | | | | Amba amba |
| | | V65 | Sun drying | Matsiroky |
| | | | | Kabiliky |
| | | | | Varilava |
| | | | | Matsiroky |
| | | | | Karara |
| | | | | Matsiroky |
| | | | | Matsiroky |

| | | | | |
|---|---|---|---|---|
| | | V66 | Sun drying | Matsiroky |
| | | | | Matsiroky |
| | | | | Varilava |
| | | | | Karara |
| | | | | Amba amba |
| | | | | Amba amba |
| | | V67 | Sun drying | Lily |
| | | | | Kabiliky |
| | | | | Varilava |
| | | | | Matsiroky |
| | | | | Karara |
| | | | | Amba amba |
| | | | | Tovy |
| | | V68 | Sun drying | Amba amba |
| | | | | Amba amba |
| | | | | Amba amba |
| | | | | Lily |
| | | | | Tovy |
| | | | | Matsiroky |
| | | | | Pelapela |
| | | | | Matsiroky |
| Ambositra | Iajaky | V69 | Sun drying | Karara |
| | | | | Tovy |
| | | | | Matsiroky |
| | | V70 | Sun drying | Matsiroky |
| | | V71 | Sun drying | Matsiroky |
| | | | | Matsiroky |
| | | V72 | Sun drying | Karara |
| | | | | Matsiroky |
| | | V73 | Sun drying | Tovy |
| | | | | Mena hariva |
| | | | | Matsiroky |
| | | V74 | Sun drying | Matsiroky |
| | | | | Tovy |
| | | V75 | Sun drying | Tovy |
| | | V76 | Sun drying | Matsiroky |
| | | V77 | Sun drying | Tovy |
| | | V78 | Sun drying | Tovy |
| | | | | Matsiroky |
| | | V79 | Sun drying | Matsiroky |
| Antsirabe | Tsena Asabotsy | V80 | Sun drying | Matsiroky |
| | | | | Amba amba |
| | | | | Amba amba |

| | | | |
|---|---|---|---|
| | | | Tovy |
| | V81 | Sun drying | Karara |
| | | | Amba amba |
| | | | Matsiroky |
| | | | Amba amba |
| | | | Tovy |
| | V82 | Sun drying | Matsiroky |
| | | | Karara |
| | | | Amba amba |
| | V83 | Sun drying | Varilava |
| | | | Mena hariva |
| | | | Matsiroky |
| | V84 | Sun drying | Karara |
| | | | Varilava |
| | | | Tovy |
| | | | Amba amba |
| | | | Matsiroky |
| | V85 | Sun drying | Matsiroky |
| | | | Tovy |
| | | | Varilava |
| | | | Ambasisy |
| | | | Lily |
| | | | Karara |
| | | | Matsiroky |
| | V86 | Sun drying | Matsiroky |
| | | | Amba amba |
| | | | Varilava |
| | V87 | Sun drying | Ambila |
| | | | Tovy |
| | | | Matsiroky |
| | | | Pelapela |
| | | | Amba amba |
| | V88 | Sun drying | Amba amba |
| | | | Matsiroky |
| | | | Matsiroky |
| | | | Tovy |
| | | | Tilapia |
| | | | Amba amba |
| | V89 | Sun drying | Varilava |
| | | | Pirina |
| | | | Pirina |
| | V90 | Sun drying | Tovy |
| | | | Varilava |

| | | | | |
|---|---|---|---|---|
| | | | | Matsiroky |
| | | V91 | Sun drying | Matsiroky |
| | | | | Varilava |
| | | | | Mena hariva |
| Antananarivo | Tsenabe Isotry | V92 | Sun drying | Varilava |
| | | | | Varilava |
| | | | | Varilava |
| | | | | Varilava mavokely |
| | | | | Mena hariva |
| | | | | Mena hariva |
| | | | | Karara |
| | | V93 | Sun drying | Vatoloha |
| | | | | Mena hariva |
| | | | Smoking | Pirina |
| | | | Sun drying | Pirina |
| | | V94 | Smoking | Tilapia |
| | | | Sun drying | Mena hariva |
| | | | Smoking | Ambasisy |
| | | V95 | Sun drying | Toho kely |
| | | | | Karara |
| | | | | Vatoloha |
| | | | | Mena hariva |
| | | | | Mena hariva |
| | | | | Mena hariva |
| | | | | Varilava |
| | | | | Karara |
| | | V96 | Sun drying | Tovy |
| | | | | Mena hariva |
| | | | Smoking | Pirina |
| | | V97 | Smoking | Toho mainty |
| | | | Sun drying | Mena hariva |
| | | | | Mena hariva |
| | | | | Mena hariva |
| | | | | Pirina |
| | | V98 | Sun drying | Matsiroky |
| | | | | Mena hariva |
| | | | | Karara |
| | | | | Pirina |
| | | | | Pirina |
| | | | | Vatoloha |
| | | | | Toho kely |
| | | V99 | Sun drying | Pirina |
| | | | | Pirina |

| | | | |
|---|---|---|---|
| | | | Mena hariva |
| | | | Karara |
| | V100 | Sun drying | Pirina |
| | | | Pirina |
| | | | Mena hariva |
| | | | Toho kely |
| | | | Vatoloha |
| Tsena Andravoahangy | V101 | Sun drying | Vatoloha |
| | | | Mena hariva |
| | | | Mena hariva |
| | | | Mena hariva |
| | | | Varilava |
| | | | Varilava |
| | V102 | Sun drying | Varilava mavokely |
| | | | Vatoloha |
| | | Smoking | Amba amba |
| | | Sun drying | Varilava |
| | | | Varilava mavokely |
| | | Sun drying | Varilava |
| | | Smoking | Amba amba |
| | | Sun drying | Varilava mavokely |
| | | | Varilava mavokely |
| | | | Varilava mavokely |
| | V103 | Sun drying | Matsiroky |
| | | | Matsiroky |
| | | | Vatoloha |
| | | | Varilava mavokely |
| | | | Varilava mavokely |
| | | | Varilava mavokely |
| | V104 | Sun drying | Varilava mavokely |
| | | | Mena hariva |
| | | | Varilava |
| | | | Varilava mavokely |
| | V105 | Sun drying | Toho kely |
| | | | Tovy |
| | | | Vatoloha |
| | | | Mena hariva |
| | | | Varilava mavokely |
| | | | Varilava mavokely |
| | | | Tovy |
| | | | Varilava mavokely |
| | | | Vatoloha |
| | | | Mena hariva |

| | | | |
|---|---|---|---|
| | | | Tovy |
| | V106 | Sun drying | Karara |
| | | Smoking | Varilava |
| | V107 | Sun drying | Pirina |
| | | | Pirina |
| | | | Varilava mavokely |
| | | | Mena hariva |
| | | | Toho kely |
| | | | Tovy |
| | | | Varilava |
| | | | Karara |

**Supplementary Table 3** Taxonomic composition and size distribution of the sampled small dried fish. For each of the ten samples, the family and scientific name of the main morpho-species identified are provided, together with the total number of individuals (N), their corresponding weight (W), and proportional biomass (P). Average standard length (SL), together with minimum and maximum values in brackets, are shown

| Sample | Family | Putative species | N | W (g) | P (%) | SL (cm) |
|---|---|---|---|---|---|---|
| **E1** | Engraulidae | *Engraulis capensis* | 199 | 68.1 | 98.6 | 4.3 [3.7-5.4] |
| | Myctophidae | *Benthosema* sp. | 4 | 0.97 | 1.4 | 3.3 [2.5-3.7] |
| **E2** | Engraulidae | *Engraulis capensis* | 67 | 36.67 | 72.3 | 5.6 [3.8-7.2] |
| | Engraulidae | *Encrasicholina punctifer* | 70 | 14.04 | 27.7 | 3.9 [3.2-4.6] |
| **E3** | Engraulidae | *Stolephorus* sp. | 235 | 47.01 | 63.6 | 4.1 [2.9-5.7] |
| | Engraulidae | *Encrasicholina pseudoheteroloba* | 49 | 8.02 | 18.6 | 4.1 [3-5.2] |
| | Clupeidae | *Sauvagella madagascariensis* | *24* | 3.25 | 10.9 | 4.3 [3.1-5] |
| | Pristigasteridae | *Pristigasteridae* sp. | *17* | 1.83 | 4.4 | 3.4 [2.4-4.3] |
| | Engraulidae | *Stolephorus indicus* | 8 | 8.02 | 2.5 | 3.4 [3.2-4.6] |
| **C1** | Clupeidae | *Spratelloides gracilis* | 154 | 63.3 | 100 | 5 [3.7-5.9] |
| **C2** | Clupeidae | *Sauvagella sp.* 1 | 509 | 53.66 | 98.4 | 3 [2.1-4.2] |
| | Cichlidae | *Oreochromis* sp. | 35 | 0.87 | 1.6 | 1.7 [1.1-1.9] |
| **C3** | Clupeidae | *Sauvagella sp.* 2 | 865 | 57.27 | 99.4 | 2.6 [2.2-3.5] |
| | Cichlidae | *Oreochromis* sp. | 3 | 0.2 | 0.3 | 1.8 [1.6-2.3] |
| | Gobiidae | *Glossogobius* sp. | 1 | 0.15 | 0.3 | 3.2 |
| **M1** | Gobiidae | - | 41 | 10.64 | 19.3 | 2.8 [1.7-5.2] |
| | Labridae | - | 10 | 8.38 | 15.2 | 4.5 [3.2-5.5] |
| | Congridae | - | 19 | 5.58 | 10.1 | 6.8 [5.7-9.2] |
| | Apogonidae | - | 19 | 5.08 | 9.2 | 2.6 [1.7-3.8] |
| | Bothidae | - | 8 | 4.62 | 8.4 | 4.2 [3.2-5.7] |
| | Scorpaenidae | - | 18 | 4.1 | 7.4 | 2.3 [1.3-4.1] |
| | Synodontidae | - | 9 | 3.32 | 6.0 | 4.6 [2.9-6.2] |
| | Pomacentridae | - | 1 | 2.87 | 5.2 | 5.6 |
| | Blenniidae | - | 5 | 1.97 | 3.6 | 3.5 [2.5-5] |
| | Acanthuridae | - | 3 | 1.37 | 2.5 | 3.3 [3.1-3.7] |
| | Scaridae | - | 1 | 1.21 | 2.2 | 5.2 |
| | Syngnathidae | - | 1 | 1.21 | 2.2 | 9 |
| | Callionymidae | - | 4 | 1.18 | 2.1 | 2.8 [1.7-5.6] |
| | Monacanthidae | - | 2 | 1.02 | 1.8 | 4.5 [3.9-5.1] |
| | Siganidae | - | 7 | 0.9 | 1.6 | 2.5 [1.7-2.7] |
| | Haemulidae | - | 2 | 0.63 | 1.1 | 3.2 [2.6-3.8] |
| | Mullidae | - | 1 | 0.39 | 0.7 | 3.9 |
| | Lethrinidae | - | 3 | 0.27 | 0.5 | 2.7 [1.4-2.7] |
| | Lutjanidae | - | 1 | 0.23 | 0.4 | 3 |
| | Chaetodontidae | - | 1 | 0.19 | 0.3 | 2.2 |

| | | | | | | |
|---|---|---|---|---|---|---|
| | Sphyraenidae | - | 1 | 0.02 | 0.04 | 2.8 |
| **M2** | Gobiidae | - | 171 | 52.7 | 84.9 | 3.7 [1.8-5.5] |
| | Callionymidae | - | 4 | 2.5 | 4.0 | 4.1 [2.8-4.9] |
| | Sillaginidae | - | 1 | 2.4 | 3.9 | 8.3 |
| | Carangidae | - | 9 | 1.67 | 2.7 | 2.9 [2-3.7] |
| | Ambassidae | - | 9 | 1.46 | 2.4 | 2.4 [2-3.1] |
| | Terapontidae | - | 2 | 0.74 | 1.2 | 3.3 [3.2-3.4] |
| | Cynoglossidae | - | 2 | 0.59 | 0.95 | 4.1 [3.7-4.5] |
| **F1** | Poeciliidae | *Gambusia holbrooki* | 2,474 | 56.33 | 93.7 | 1.5 [1.1-2.8] |
| | Cichlidae | *Coptodon zillii* | 56 | 3.77 | 6.3 | 1.5 [0.9-3.1] |
| **F2** | Eleotridae | *Eleotris* sp. 1 | 333 | 50.84 | 85.8 | 3.2 [2.5-5.2] |
| | Poeciliidae | *Xiphophorus hellerii* | 61 | 8.12 | 13.7 | 2 [1.4-3.2] |
| | Cichlidae | *Oreochromis* sp. 1 | 3 | 0.3 | 0.5 | 2 [1.4-2.7] |
